# Special Issue: Demographic and Mental Health Assessments in the Adolescent Brain and Cognitive Development Study: Updates and Longitudinal Trajectories

**DOI:** 10.1101/2021.07.05.21260023

**Authors:** Deanna M. Barch, Matthew D. Albaugh, Arielle Baskin-Sommers, Brittany E. Bryant, Duncan B. Clark, Anthony Steven Dick, Eric Feczko, John J. Foxe, Dylan G. Gee, Jay Giedd, Meyer D. Glantz, James J. Hudziak, Nicole R. Karcher, Kimberly LeBlanc, Melanie Maddox, Erin C. McGlade, Carrie Mulford, Bonnie J. Nagel, Gretchen Neigh, Clare E Palmer, Alexandra S. Potter, Kenneth J. Sher, Susan F. Tapert, Wesley K. Thompson, Laili Xie

## Abstract

The Adolescent Brain Cognitive Development (ABCD) Study of 11,880 youth incorporates a comprehensive range of measures assessing predictors and outcomes related to mental health across childhood and adolescence in participating youth, as well as information about family mental health history. We have previously described the logic and content of the mental health assessment battery at Baseline and Year 1. Here, we describe changes to that battery and issues and clarifications that have emerged, as well as additions to the mental health battery at the 2-, 3-, 4-, and 5-year follow-ups. We capitalize on the recent release of longitudinal data for caregiver and youth report of mental health data to evaluate trajectories of dimensions of psychopathology as a function of demographic factors. For both caregiver and self-reported mental health symptoms, males showed age-related decreases in internalizing and externalizing symptoms, while females showed an increase in internalizing symptoms with age. Multiple indicators of socioeconomic status (caregiver education, family income, financial adversity, neighborhood poverty) accounted for unique variance in both caregiver and youth-reported externalizing and internalizing symptoms. These data highlight the importance of examining developmental trajectories of mental health as a function of key factors such as sex and socioeconomic environment.

---

As described in numerous publications to date, the Adolescent Brain Cognitive Development□(ABCD) Study is a unique longitudinal study of almost 12,000 youth in the United States that will inform our understanding of the environmental, genetic, neurobiological, and behavioral factors that promote health and well-being, as well as those that put youth at risk for challenges in adaptive functioning and mental health. The ABCD Study® started when youth were 9 and 10 years old, and will run for at least 10 years. As described previously (Barch et al. 2018; Casey et al. 2018; Iacono et al. 2018; Luciana et al. 2018; Hagler et al. 2019; Karcher and Barch 2020) and in other papers in this special issue, the ABCD Study is collecting a large range of data on each youth, including physical health data, neuroimaging data, biomarkers (e.g., hormones, DNA), assessment of cognitive function, reported substance exposures and use, as well as measures of the youth’s family and environment, including measures of the youth’s neighborhood and societal environment. As a central part of understanding well-being in youth, the ABCD Study assesses a broad range of constructs that both predict and denote outcomes related to adaptive function and mental health. In a previous publication (Barch et al. 2018), we described the process and principles that drove the development of the baseline battery of mental health-relevant measures. Here, we describe updates and changes to the mental health battery that have been or are being incorporated over the first six in-person assessment waves of the study, the principles that are driving decisions about what assessments are added to the study, which measures will continue to be assessed and at what frequency, and which measures have been dropped. We also overview known issues or considerations with measures that are being collected, and provide new data on longitudinal trajectories of both parent and youth reported mental health in relation to a number of important demographic factors (sex, race, parental education, socioeconomic status). The current membership of the ABCD Mental Health Assessment Workgroup is shown in Table S1.

## 1. Brief Overview of the Baseline ABCD Demographics Mental Health Battery

We described the justification and purpose of the baseline ABCD baseline demographics and mental health battery in the first ABCD Special Issue in Development Cognitive Neuroscience. The development of this battery was guided by the existing literature in relevant constructs and measures, developmental appropriateness and ability to be usable across adolescence, the feasibility and reliability of use across many sites, evidence about the psychometric properties of the instruments, ability to harmonize with previous or ongoing studies where possible, and measures that have been recommended as common data elements by the PhenX initiative (Stover et al. 2010; Hamilton et al. 2011; Maiese et al. 2013; McCarty et al. 2014) or other NIH assessment initiatives (Conway et al. 2014; Barch et al. 2016). These same principles continued to guide our decisions about updates and expansions to the ABCD Mental Health Battery over the ongoing waves of assessment.

As shown in Tables 1 and 2, we assess constructs relevant to the youth’s adaptive function and mental health from the perspective of the caregiver, the youth themselves, and the youth’s teacher. In terms of demographics, we collect a range of information about socioeconomic status and financial insecurity (Diemer et al. 2012), household composition (McLoyd 1998; Smith et al. 2015), race and ethnicity of the youth and the extended family, gender identity and sexual orientation. Since the publication of the original paper on the mental health assessments in the ABCD, Gender Identity and Sexual Health has become a stand-alone workgroup, and thus the changes to the measures of gender identity and sexual orientation are described in another paper in this special issue and modifications to the demographic measures over assessment waves are described in Section 2 below. We also ask both youth and parents to report on school performance, numbers of friends and friendship quality and bullying (see Section 6 for additional measures of peer relationships (Prinstein et al. 2001; De Los Reyes and Prinstein 2004) and cyberbullying (Stewart et al. 2014) added in Year 2).

**Table 1:**
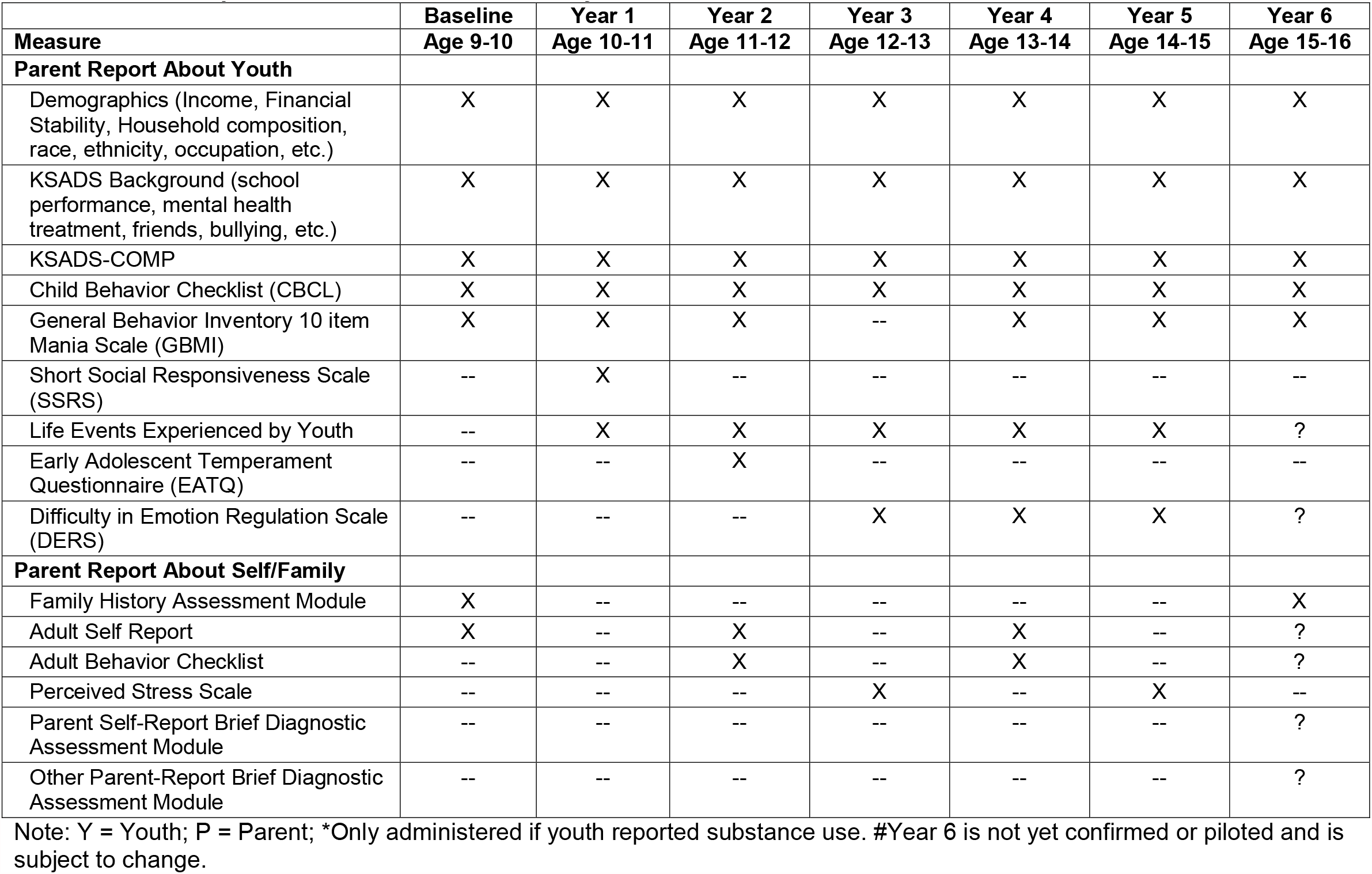
Parent Report About Youth and Self/Family.

**Table 2:** Youth Report About Self.

| **Measure** | **Baseline**<br>**Age 9-10** | **Year 1**<br>**Age 10-11** | **Year 2**<br>**Age 11-12** | **Year 3**<br>**Age 12-13** | **Year 4**<br>**Age 13-14** | **Year 5**<br>**Age 14-15** | **Year 6#**<br>**Age 15-16** |
| --- | --- | --- | --- | --- | --- | --- | --- |
| KSADS Background (school performance, mental health treatment, friends, bullying, etc.) | X | X | X | X | X | X | X |
| KSADS-COMP (Kobak et al. 2013) | X | X | X | X | X | X | X |
| Brief Problem Monitor-Youth (BPM-Y) | -- | X | X | X | X | X | ? |
| NIH Toolbox Positive Affect Items | -- | X | -- | X | -- | X | -- |
| 7-Up Mania Scale (7-UP) | -- | X | -- | X | -- | X | -- |
| Psychosis Questionnaire-Brief Child (PQ-BC) | X | X | X | X | X | X | X |
| Urgency, Perseverance, Premeditation, and Sensation Seeking (UPPS-Child) | X | -- | X | -- | X | -- | ? |
| Behavioral Inhibition and Behavioral Activation Scale (BIS-BAS) | X | -- | X | -- | X | -- | ? |
| 10-Item Delinquency Scale | -- | X | X | X | X | X | X |
| Life Events Experienced by Youth | -- | X | X | X | X | X | X |
| Emotion Regulation Questionnaire | -- | -- | -- | X | X | X | X |
| **Number of Friends and Close Friends** |  |  |  |  |  |  |  |
| Peer Experiences Questionnaire | -- | -- | X | X | X | X | X |
| Cyberbullying Questionnaire | -- | -- | X | X | X | X | X |

#Year 6 is not yet confirmed or piloted and is subject to change.

### Mental Health Diagnoses

From parent and youth perspective, information is obtained about both current and lifetime mental health diagnoses of the youth using a validated and computerized *Kiddie Schedule for Affective Disorders and Schizophrenia (KSADS) for DSM-5 (KSADS-COMP)*, developed by Dr. Joan Kaufman and Dr. Ken Kobak with NIH SBIR support (Kobak et al. 2013; Kobak and Kaufman 2015). This is a self-administered, computerized version that does not involve a clinician for either the parent or the youth, though the youth are supported in completing the KSADS-COMP by trained research assistants. In Section 3 below, we provide more information about changes in this measure over assessment waves and known issues or considerations in the use of data from the KSADS-COMP.

### Dimensional Measures of Mental Health Relevant Constructs

Dimensional measures of mental health from parents, youth and teachers were also obtained. As shown in Table 1, starting at baseline, parents report annually about the youth’s behavior using the *Child Behavior Checklist* (CBCL) (Achenbach 2009) as a broad measure of many domains and about dimensional mania symptoms using the ten-item Mania Scale (Youngstrom et al. 2008) derived from the 73-item Parent General Behavior Inventory (PGBI) for Children and Adolescents (Youngstrom et al. 2001). As shown in Table 2, youth reported about their own mental health every six months (starting at the first six month mid-year assessment) using the Brief Problem Monitor for youth (BPM-Y) (Achenbach 2009) and the positive affect items from the NIH Toolbox Battery (Gershon et al. 2013; Salsman et al. 2013). In Section 4 below, plans to move the assessment from the BPM-Y to the more comprehensive *Youth Self-Report* (YSR) are outlined. Youth also report annually on psychotic-like experiences using the Prodromal Questionnaire Brief Version-Child (Karcher et al. 2018; Karcher et al. 2020), and bi-annually on impulsivity using a brief urgency, perseverance, premeditation, and sensation seeking (UPPS) scale (Watts et al. 2020), mania using the 7-item child report of mania called the 7-Up (Youngstrom et al. 2013) (starting at Year 1), and on behavioral activation and inhibition using the BIS-BAS (Carver and White 1994; Pagliaccio et al. 2016).

At the Year 1 assessment, as shown in Tables 1 and 2, parents also started reporting on behaviors relevant to the autism spectrum, using the brief Social Responsiveness Scale (Reiersen et al. 2008). We also started asking youth at Year 1 to annually report on their own delinquency-relevant behaviors using a 10-item shortened version of the scale developed for use in the Causes and Correlates of Delinquency Program (Hoeve et al. 2008; Theobald et al. 2014). See Section 5 for issues related to the use and interpretation of data from this measure. As described in more detail in Section 9, at Year 1 we also began annual administration of the *Adverse Life Events Scale* (Tiet et al. 2001; Grant et al. 2004) from the PhenX collection asking for both parent and youth reports about events that the youth experienced. At Year 2, we also added a parent report measure of youth temperament called the Early Adolescent Temperament Scale (Latham et al. 2020) at Year 2 (see Table 1 and Section 8) and both parent (Difficulties in Emotion Regulation Scale (Bardeen et al. 2016; Lee et al. 2016; Benfer et al. 2019; Bunford et al. 2020) and youth (Emotion Regulation Questionnaire (Gullone and Taffe 2012)) report measures of emotion regulation starting at Year 3 (see Tables 1 and 2 and Section 7).

### Teacher Report

To provide converging evidence about the youth’s behavior, families are asked to give permission to allow their youth’s teacher to complete the Brief Problem Monitor – Teacher Form (Achenbach 2009) at each assessment wave. See Section 4 for discussion of how teachers were selected and considerations in the use of BPM-T data.

### Parent/Family Mental Health and Personality

As shown in Table 1, at baseline we used a version of the Family History Assessment Module Screener (FHAM-S) (Rice et al. 1995) that was used in the National Consortium on Alcohol and Neurodevelopment in Adolescence (NCANDA) study (http://www.ncanda.org/index.php) (Brown et al. 2015). The engaged parent/caretaker reports on the presence/absence of symptoms associated with alcohol use disorder, substance use disorder, depression, mania, psychosis, and antisocial personality disorder in all 1^st^ and 2^nd^ degree “blood relatives” of the youth, defined as biological relatives including full and half-siblings, parents, grandparents, and aunts and uncles. See Section 10 for more information on the scoring of the FHAM-S data and interpretation. We also ask the study-engaged caretaker to complete the Adult Self Report (Achenbach 2009) bi-annually, and to also complete the *Adult Behavior Checklist* (Achenbach 2009) about the other caregiver bi-annually starting at Year 2. See Section 4 on how the other parent was selected and interpretation of these data. We began asking the study-engaged caretaker to complete the Perceived Stress Scale (Cohen et al. 1983) about their own level of stress bi-annually starting at Year 3 (see Section 11). Ideally, it is the same caretaker every year, but this has not always been the case. A more detailed update on parent mental health history (both engaged caretaker and report on other parent) is planned for Year 6 (see Section 10)

## 2. Updates to Demographic Assessments in the ABCD Study

Caregivers complete demographic assessments at every visit. Every year, caregivers provide demographic information about the child, themself and a partner, which specifically refers ‘to any significant person in [the] child’s life that helps in raising [the] child or has helped for more than 2 years and who is involved in 40% or more of the daily activities of [the] child’. Information about both the caregiver and child includes age, sex, race, ethnicity, gender identity, religious preferences and experiences and native language. The caregiver additionally provides information about their current marital status, highest education level, current employment status, work sector, income, and their partner’s relationship to child, highest education level, employment status, work sector and income, and a combined total household income. In addition, every year caregivers also report how much time the child spends in different households, how people live at their address, household member’s relationship to the caregiver and details of any financial difficulties faced in the past 12 months (e.g. could not afford food, had services turned off because payments were not made). At Year 1 (and continuing annually), additional questions were introduced regarding whether the child was covered by any health insurance or coverage plans at the time of data collection. At Year 2 (and continuing annually), an occupation survey was added to provide a detailed characterization of both the caregiver and partner’s work sector and job title to allow a more precise quantification of socioeconomic status (SES).

## 3. KSADS-COMP for DSM 5

Table S2 shows the KSADS modules completed by parents and youth at each annual assessment There are a number of modules that parents have reported on every year (psychosis, eating disorders, ADHD, conduct disorder) since we did not have other assessments of these constructs. Most of the other modules are completed every other year by parents. For youth, the only module they complete every year is the suicidality module, as this is the only self-report of this information, with mood disorders, social anxiety, generalized anxiety and sleep every other year starting at baseline, and eating disorders and conduct disorder added at year two. Youth complete the alcohol or drug use modules starting in Year 1 if they report use of any substances. As can be seen in Table S2, parents are completing many more modules than youth, particularly at the early assessment waves. This was purposeful, both from the perspective of burden on the youth (they have many assessments to complete) and because we felt that there were many domains on which parents would be much better reporters than children (e.g., externalizing disorders, tics) earlier in adolescence. However, we have been gradually adding more modules for youth to complete over the assessment waves, as we feel they are better able to accurately report on facets of their behavior. The first three years of assessments (baseline, Year 1, Year 2) used the KSADS 1.0, which was the version available at the time that the study started. We switched to the KSADS 2.0 at Year 3 as this updated version incorporates several improved features, including better assessments of Autism Spectrum and Psychotic disorders.

There are several important issues that inform the use of the KSADS data. First, the KSADS-COMP was designed to be an efficient self-administered evaluation of DSM symptoms, therefore, response options across the current (past two-weeks) and past (ever) questions do not necessarily correspond to recommended criterion thresholds for DSM symptoms. For example, the questions framed as ever experiencing a symptom or displaying a behavior are evaluated with a two-choice (yes/no) option. For disorders, such as Conduct Disorder or Major Depressive Disorder, where the behaviors should occur repeatedly or over a certain number of days to meet the symptom criterion, the KSADS-COMP diagnosis is most conservatively conceptualized as an estimated or approximated DSM diagnosis. Second, while there is an item labeled “Diagnosis - Other Specified Neurodevelopmental Disorder, Autism Spectrum Disorder, full criteria not assessed (F88.0)” this should not be used as an indication of an Autism Spectrum Disorder. The KSADS 1.0 asks questions about some behavioral features relevant to Autism Spectrum Disorder, but does not do a full evaluation nor does it generate a diagnosis. Third, there are diagnoses for Schizophrenia, Schizophreniform and Schizoaffective disorder in KSADS 1.0, but they were not based on a full evaluation and thus should not be used as indicators of a diagnosis. Fourth, the Agoraphobia questions were accidentally omitted when we changed platforms for a brief period of time (6 months). Fifth, all of the diagnoses and symptoms are presented as 0 (absent) or 1 (present), and do not use the more elaborated coding typically used in the KSADS (0 = not enough information, 1 = absent, 2 = subthreshold, 3 = present), though we are working on generating these data for future releases. Sixth, the KSADS uses a screening module, and then additional questions are asked if items on the screening module are answered affirmatively. For symptom items, we coded them separately if they were deliberately not asked (888) because the individual did not answer screening items affirmatively (i.e., due to branching logic), as compared to not administered at that wave (555). Sixth, the computerized KSADS 1.0 shows higher than expected rates of caregiver- and youth-reported past manic episodes (i.e., the caregiver-reported prevalence of Bipolar I Disorder, most recent past episode manic is 2.6%). To address for these higher than expected rates, we recommend rescoring Bipolar I Disorder, most recent past episode manic so that the youth has to meet criteria for past manic episode and any current or past depressive disorder (e.g., major depressive disorder, persistent depressive disorder, other specified depressive disorder) in order to meet diagnostic criteria.

Lastly, in May of 2021, the KSADS-COMP originators did a review of diagnostic criteria used in the programming algorithms, and found several errors that likely let to overestimates of diagnoses. The needed modification are: a) need to include impairment in the diagnostic criteria for Major Depression and Persistent Depressive Disorder; b) need to include onset before age 10 in the diagnostic criteria for Disruptive Mood Dysregulation Disorders; c) need to require impairment in two domains, not just one, for Attention Deficit Hyperactivity Disorder; d) need to require an illness duration of 6 months or more for Agoraphobia; e) need to include an illness duration of three months or longer for Anorexia; and f) need to include the presence of all three criterion A symptoms for Autism (relevant to the 2.0 version). The KSADS-COMP will be modified with these updated criteria by June 1 of 2021, and the ABCD team is working with KSADS-COMP to correct diagnoses in already acquired data.

## 4. ASEBA Scales in the ABCD: BPM-Y, BPM-T, YSR, ASR and ABCL

The Achenbach System of Empirically Based Assessment (ASEBA) was employed in order to obtain longitudinal, dimensional assessments of child and parent/caregiver psychopathology. This family of instruments assesses a number of empirically derived syndromes that were developed in a “bottom up” fashion using factor analytic methods (Achenbach and Rescorla 2001). The structure of these empirically derived syndromes are remarkably stable across various societies and cultures—making it an ideal set of instruments given ABCD’s large and diverse sample (Achenbach and Rescorla 2007; Achenbach and Rescorla 2015). ASEBA empirically derived syndromes can be assessed across the developmental span, allowing for the same psychopathological constructs to be measured longitudinally across ABCD’s 10-year duration. The ASEBA battery has been used in other large longitudinal studies such as Generation R study in the Netherlands (Jaddoe et al. 2006), which will allow for cross-cultural comparison and facilitate communication of findings.

The ASEBA family of instruments affords for multi-informant assessment of developmental psychopathology. This methodological feature is capitalized upon in the ABCD Study, enabling multiple informants to rate child behavior in different settings (e.g., home, school). Starting at the baseline visit, the parent/caregiver annually completes the Child Behavior Checklist (CBCL) on their child’s behavior. Beginning at the 6-month follow-up, the youth version of the Brief Problem Monitor (BPM-Y) is administered every 6 months. Thus, the BPM-Y is particularly useful in monitoring the longitudinal course of youth functioning. Complementing parent- and self-report, teachers complete the Brief Problem Monitor teacher’s form (BPM-T) on youth. In selecting the BPM-T informant, caregivers were asked to choose the teacher who had the most frequent contact with their child (preferably not Gym/Physical Education teachers unless there were no academic teachers available). An email was then sent to the identified teacher which included a confidential link to the web-based BPM-T. If a child had a study visit during the summer months, study staff would wait to gather teacher information at the 6-month follow-up phone call. This was done to ensure that the teacher completing the BPM-T was familiar with the child. To date, 42%, 54%, and 54% of teachers have provided reports for Baseline, Year 1, and Year 2 respectively.

The BPM-Y and BPM-T were administered in lieu of the longer Youth Self-Report (YSR) and Teacher’s Report Form (TRF) measures (Achenbach and Rescorla 2001; Achenbach et al. 2017) to lessen the burden on participants and teachers. The BPM-Y and BPM-T consist of a subset of items from the YSR and TRF (19 and 18 items, respectively) and yield scores for Internalizing, Attention Problems, Externalizing, and Total Problems scales (Achenbach et al. 2017), though they do not address psychosis. The YSR will be administered mid-year during Year 4 which will provide comprehensive coverage of all ASEBA empirically derived syndromes. Lastly, parent/caregiver functioning is a vital component of the family environment.

Conveniently, ASEBA allows for the same empirically derived psychopathological constructs to be assessed in parents/caregivers. Beginning at the baseline visit, the parent/caregiver annually completes the Adult Self Report (ASR) on their own behavior (Achenbach and Rescorla 2003). At Year 2, the parent/caregiver also completes the Adult Behavior Checklist (ABCL)(Achenbach and Rescorla 2003) on the other biological parent who lives with the child, or on the other parent/caregiver in the home who has lived with the child for at least 6 months. If the primary parent/caregiver did not live with the child’s other parent/caregiver, the ABCL was still completed provided the child had regular contact with the other parent/caregiver.

## 5. Brief Delinquency Measure

While the baseline ABCD protocol includes some items in which parents reported on youth delinquent behaviors (e.g., KSADS conduct disorder), youth self-report in this arena was not included. “Delinquent behaviors” may be defined as antisocial acts that violate societal norms and laws (Isen et al. in press). Since youth may engage in delinquent acts (i.e., violating social norms and laws:(Isen et al. in press)) that escape detection by parents and may not be recorded in official records, self-reported delinquency has become central to the study of adolescent delinquent and/or criminal behavior (Piquero et al. 2002). To provide a brief assessment of a range of delinquent behaviors varying in severity, ten items were selected from a version of the Self-Reported Delinquency Scale (SRD) (Elliot et al. 1989) adapted for the Pittsburgh Youth Study and the Pittsburgh Girls Study (Loeber et al. 2008). The ten items, titled the Brief Delinquency Measure, were added starting in Year 1 (see Clark et al. Table 1 for items).

In recent work, we have conducted analyses of measurement invariance across race and sex and examined the items for differential item functioning. These analyses, which are reported in detail in Clark et al. (Clark et al. in preparation) revealed evidence of differential item function. The item related to being arrested by the police showed that, for the same putative level of trait delinquency, black youth were much more likely to have been arrested by police. This pattern is consistent with the evidence of systemic racism in regards to black youths experience with police. As such, we are evaluating whether to move the item in regards to being arrested to a new measure that asks more extensive questions about the role of police in a child’s community to assess this information in a different context. Further, other items asked about youth being told they are being too rowdy, which may also reflect differential experiences of black youth (Fadus et al. 2020). This general review of measures also revealed concerns that this 10-item measure may need to be modified or supplemented by items from other scales to more fully capture a broader range of delinquent behaviors in a less biased manner. Thus, the measures of delinquency and associated antisocial traits are being reexamined to reduce redundancy, provide appropriate coverage across this diverse sample, and to ensure that items potentially reflecting others’ reactions to youth that may reflect implicit or explicit bias are recognized as environmental reactions to behaviors and not necessarily used as indicators of problem behavior on the part of the youth.

## 6. Peer Relationships and Cyberbullying

Peer relationships are a key part of adolescent life. Peer victimization (both being a perpetrator and/or victim of bullying) has negative associations with outcomes that last well beyond adolescence. To capture these experiences, beginning in Year 2, youth complete the Revised Peer Experiences Questionnaire (Prinstein et al. 2001; De Los Reyes and Prinstein 2004). This is an 18-item questionnaire that yields scores for overt, relational and reputational aggression (both being a victim and perpetrator). Adolescent peer relationships occur both in person and virtually, and so two questions were added to capture cyberbullying (Stewart et al. 2014). Youth are asked “Have you ever been cyberbullied, where someone was trying on purpose to harm you or be mean to you online, in texts, or group texts, or on social media (like Instagram or Snapchat)?” They are also asked about cyberbullying another person.

## 7. Emotion Regulation

Emotion regulation, or the ability to modulate one’s emotions, is important for healthy functioning and undergoes substantial change across development (Cole et al. 1994).

Moreover, difficulties with emotion regulation have been identified as a transdiagnostic risk factor for many forms of psychopathology (Southam-Gerow and Kendall 2002; Aldao et al. 2016). To assess emotion regulation in the ABCD Study, we added the child-reported Emotion Regulation Questionnaire for Children and Adolescents (ERQ-CA; (Gullone and Taffe 2012)) and parent-reported Difficulties in Emotion Regulation Scale (DERS-P; (Bunford et al. 2020)). The ERQ-CA, which was developed based on the original Emotion Regulation Questionnaire (Gross and John 2003), examines the tendency to use two specific emotion regulatory strategies—cognitive reappraisal and expressive suppression. We selected 3 items assessing cognitive reappraisal (e.g., “When I want to feel less bad (e.g., sad, angry, worried) about something, I think about something different”) and 3 items assessing expressive suppression (e.g., “I control my feelings by not showing them”) from the 10-item measure. The ERQ-CA has shown good internal consistency and test-retest reliability over 12 months in a sample of 10 to 18 year-olds. The DERS-P examines difficulties across domains related to nonacceptance, goals, impulses, strategies, awareness, and clarity. The DERS-P was validated in a sample of 11 to 17 year-olds and has been shown to have good concurrent and convergent validity (Bunford et al. 2020). We selected 29 items from the DERS-P, eliminating 7 items administered in the original study that did not load onto any factors.

## 8. Temperament and Personality

In Year 2, the Early Adolescent Temperament Questionnaire-Revised (EATQ-R; parent version)(Capaldi and Rothbart 1992) was added to the parent assessment battery. The EATQ-R, designed to assess temperament in adolescents from ages 9-15, assesses eight primary dimensions of temperament, two dimensions of behavior, and three higher-order dimensions that subsume these primary dimensions. The eight primary temperament dimensions include: Activation Control-capacity to perform an action when there is a strong tendency to avoid it, Affiliation-desire for warmth and closeness with others, independent of shyness or extraversion, Attention-capacity to focus attention as well as to shift attention when desired, Fear-unpleasant affect related to anticipation of distress, Frustration-negative affect related to interruption of ongoing tasks or goal blocking, Surgency/High Intensity Pleasure-pleasure derived from activities involving high intensity or novelty, Inhibitory Control-capacity to plan, and to suppress inappropriate responses, and Shyness-behavioral inhibition to novelty and challenge, especially social. The two behavioral scales include: Aggression-hostile and aggressive actions (person & object directed physical violence, direct & indirect verbal aggression, hostile reactivity) and Depressive Mood-unpleasant affect and lowered mood, loss of enjoyment and interest in activities.

Inclusion of parent-rated, dimensional, temperamental traits is useful for obtaining perceptions of individual differences from the perspective of an adult who interacts considerably with the youth. Future personality assessments will maintain the focus on impulsivity-related traits given their relevance for understanding substance use disorders and other externalizing psychopathology, and on behavioral inhibition for understanding internalizing disorders. In future waves, we plan to assess youth-reported “Big Five” traits to have a well-established framework for characterizing personality more generally and track personality development into late adolescence and young adulthood.

## 9. Life Events

The original Adverse Life Events Scale (Tiet et al. 2001) is a 25-item questionnaire that includes a variety of events such as whether parent(s) had drug problems, lost a job, went to jail, was away from home more than usual, left home/divorced; family member or close friend was injured, seriously sick, or died; participant was seriously sick or injured; or participant saw a crime/accident or was the victim of a crime. Updates to the protocol at Year 4 included items for parent/caregiver being deported; youth being placed in foster care; seeing someone getting beaten up in school/neighborhood or shot at; and having a lockdown at school due to concerns about violence. At the yearly study visit, the youth and parents indicate whether each life event happened to them in the prior year (yes/no). For all events that did happen, parents and youth are asked whether the experience was good or bad (mostly good, mostly bad, not applicable, or don’t know). They are then asked how much the event affected them (not at all, a little, some, or a lot). Scoring yields the number of total events; events characterized by the participant as bad (response of mostly bad and a little, some or a lot bad); and events characterized by the participant as good (response of mostly good and a little, some or a lot good).

## 10. Family History

As described previously, in ABCD we employed a version of the Family History Assessment Module Screener (FHAM-S) (Rice et al. 1995) that was used in the National Consortium on Alcohol and Neurodevelopment in Adolescence (NCANDA) study (http://www.ncanda.org/index.php). In the ABCD FHAM-S version, we had a caregiver report on the presence/absence of symptoms associated with alcohol use disorder, substance use disorder, depression, mania, psychosis, and antisocial personality disorder in all 1st and 2nd degree “blood relatives” of the youth. (That is, biological relatives including full and half-siblings, parents, grandparents, and aunts and uncles.) Note, however, that these assessments are quite abbreviated. Still, assessing each participant’s pedigree in this way allowed us to characterize not only the family history of each participant with respect to each of the classes of disorder listed above but also to create alternative indices beyond simple global designations such as the presence or absence of a family history of a given disorder. Because of complexities in scoring the interview, our data release now includes summary variables of reported parental disorders (e.g., mother only, father only, both father and mother) for each of the classes of disorders assessed. Users can derive more complex pedigree measures (e.g., multigenerational typologies, family history density measures) that includes measures ranging from *continuous indices* of genetic risk such as family history density that considers the number of affected 1st and 2nd degree relatives in the pedigree (Stoltenberg et al. 1998) whether or not the family history is unilineal or bilineal (i.e., matrilineal, patrilineal, or both) (Volicer et al. 1983) or unigenerational (parental generation only) or multigenerational (i.e., parent and grandparent on one side) (Finn et al. 1990). However, because of the various permutations of such approaches, users must create their own based on the needs of their own particular study.

Examination of the frequencies of reported parental alcohol problems, drug problems, conduct/antisocial problems, problems associated with “nervousness”, mania, psychotic symptoms, suicidality, professional help-seeking, and inpatient hospitalization yielded overall prevalence and sex differences consistent with expectation. Aggregating across different conditions (e.g., creating a history of parental externalizing disorders by aggregating across substance use disorders and antisocial behavior, internalizing by aggregating “nerves” and depression, and thought problems by aggregating across mania and psychotic symptoms) showed higher prevalence than their constituent conditions. Further aggregation, combining externalizing, internalizing, and thought problems yielded prevalence indicating that nearly half of the ABCD families (47%) reported one or both biological parents to have been affected by one or more of the conditions assessed.

At the time of baseline assessment, some members of a pedigree had yet to enter their period of risk or were still transitioning through early stages of their period of risk for the disorders being assessed. Additionally, informants may only become aware of a problem in a relative subsequent to the baseline assessment. Consequently, reassessment of family history is planned for a future follow-up, most likely the Year 5 or 6 follow-up. It has not yet been determined if the same family history interview will be readministered (which has the virtue of maintaining the same method) or will be replaced by a more complete assessment that comports better with contemporary diagnostic constructs. In addition, in the course of collecting the family history data we discovered that the interview program was not robust to fully assessing some large pedigrees (e.g., no more than five maternal aunts could be assessed but some informants indicated 6 or more maternal aunts). In the next follow-up, no constraints will be imposed on pedigree size. While the planned re-administration will strengthen the assessment of family history beyond the baseline assessment, the assessment will nonetheless remain somewhat crude owing to the burden of more complete assessment of all 1st and 2nd degree relatives and users should be mindful of the fact that family history methodology is known to have relatively low sensitivity.

## 11. Parental Perceived Stress

A relationship between perceived stress and behavioral and health outcomes is well established (DeVries et al. 2007; Mukhara et al. 2018; Turkson et al. 2019). The Perceived Stress Scale (PSS) was first published by Cohen and colleagues in 1983 (Cohen et al. 1983). The PSS is an accessible and effective metric of perceived stress, and has demonstrated utility in understanding the relationship between stress and a myriad of both behavioral and health endpoints (Golden-Kreutz et al. 2005; Robles et al. 2016; Whitehead and Bergeman 2017; Barutcu Atas et al. 2021). In addition to predictive value for outcomes specific to the individual completing the PSS, this metric has also proven useful in understanding the role of parental perceived stress in adolescent-related outcomes (Slaughter et al. 2020; Koning et al. 2021; Tara et al. 2021). The role of parental behaviors and parental perceived stress are salient influences during adolescence; therefore, a PSS assessment that parents complete about their own stress has been added to the collected metrics.

## 12. Trajectories of Parent and Child-Report Mental Health

As described above, one of the benefits of the ABCD Study is the longitudinal assessments of both parent and child reported mental health in a large non-treatment seeking sample. There have been several previous studies using epidemiological or non-treatment seeking samples that have reported data on longitudinal trajectories of mental health, as well as the relationship of various demographic factors relevant to understanding mental health among youth. In terms of age-related differences in mental health, there is consistent evidence from studies in both the United States, Canada and Europe that levels of depression tend to increase from school age into adolescence (Strohschein 2005; Van Oort et al. 2009; Robbers et al. 2010; Ormel et al. 2012; Ferro et al. 2015; Coley et al. 2019; Antolin-Suarez et al. 2020), with evidence that this increase is greater in females than males (Bongers et al. 2003). Patterns for anxiety are more mixed, with some evidence for decreases in various forms of anxiety from school age to adolescence (Van Oort et al. 2009; Ormel et al. 2012).In general, the data suggest that aggressive, attentional and rule-breaking problems tend to decrease from middle childhood to adolescence (Bongers et al. 2003; Strohschein 2005; Fanti and Henrich 2010; Robbers et al. 2010), but with some exceptions (Keiley et al. 2000; Ormel et al. 2012). In terms of sex relationships, evidence consistently shows higher rates of depression among females (Faravelli et al. 2013), typically diverging at early adolescence (Bongers et al. 2003; Brown et al. 2007; Robbers et al. 2010). In contrast, rates of anxiety show less evidence of sex differences in childhood/adolescence (Faravelli et al. 2013), and rates of externalizing problems tend to be higher in boys across childhood and adolescence (Robbers et al. 2010; Robbers et al. 2011), though with some evidence for convergence in later adolescence (Bongers et al. 2003).

The evidence about differences in levels of depression, anxiety, and externalizing symptoms in childhood and adolescence as a function of race or ethnicity is more mixed. In the Monitoring the Future study, Black students reported lower symptoms of depression in 10^th^ and 12^th^ grade compared to White students, but other students of color reported higher levels of depression (Coley et al. 2019). In the National Longitudinal Study of Youth, depression was lower among non-Hispanic Black youth than white youth In the National Longitudinal Study of Youth, non-Hispanic Black youth reported less depression than White youth (Strohschein 2005). In the National Longitudinal Study of Adolescent to Adult Health, Hispanic and Asian youth showed the highest levels of depression and white youth the lowest, with black youth falling in between (Brown et al. 2007). In the National Comorbidity Survey Replication-Adolescent supplement, Hispanic youth and non-Hispanic black youth had higher rates of mood and anxiety disorders than non-Hispanic white youth (Georgiades et al. 2018). However, these differences were eliminated when sociodemographic factors including parental education and family income were considered (Georgiades et al. 2018). In the National Survey of Children’s Health from 2003, 2007, and 2011, rates of attention deficit disorder symptoms were highest among White children (Collins and Cleary 2016). In the National Longitudinal Study of Youth, conduct problems were higher among non-Hispanic black youth than white youth, but it was not clear if these race-related differences were eliminated or reduced with socioeconomic factors were considered (Strohschein 2005). In contrast, the National Comorbidity Survey Replication-Adolescent supplement, non-Hispanic black youth and Asian youth had lower rates of behavior disorders than non-Hispanic white youth (Georgiades et al. 2018). Critically, few of these studies address critical contextual factors that are often confounded with race and ethnicity, including factors such as socio-economic status of families and neighborhoods, or other stressors such as discrimination experiences and the effects of systemic racism, all of which likely influence mental health.

Apropos the concerns about contextual factors often confounded with race and ethnicity, the evidence in regards to the relationship between socioeconomic status (SES) and mental health in children and adolescents is robust, with youth living in lower SES households showing consistently higher rates of depression, anxiety, and externalizing symptoms in both the United States and Europe (Goodman et al. 2003; Strohschein 2005; Amone-P’Olak et al. 2009; Robbers et al. 2010; Letourneau et al. 2011; van Oort et al. 2011; Reiss 2013; Coley et al. 2019; Antolin-Suarez et al. 2020). However, what is less clear is whether different facets of SES relate differentially to youth mental health, such as family income, caregiver education, indices of financial insecurity or neighborhood SES (Denny et al. 2016; Coley et al. 2019).

### Current Analyses

We used the data from the most recent ABCD data release to characterize the trajectories of both parent and child reported internalizing (depression and anxiety) and externalizing (ADHD, oppositional defiant disorder, conduct disorder) symptoms between the ages of 9 and 13 as a function of demographic features. Based on the existing literature, we expected both parent and child reports of internalizing symptoms to increase across the course of development, particularly among girls. In regards to externalizing symptoms, we expected rates to be higher in boys and to decrease as a function of age. In regards to race and ethnicity, we did not have strong a priori hypotheses, as we conducted these analyses including a variety of SES related factors that may have influenced race/ethnicity related differences in mental health in prior studies. Based on the existing literature, we expected that lower socioeconomic status (lower caregiver education, lower family income, greater financial insecurity) would be associated with overall higher reports of internalizing and externalizing symptoms. However, we did not have strong a priori hypotheses as to whether the different indicators of SES would relate differentially to child mental health.

## Methods

### Participants

Data were from caregiver and youth participants from the current ABCD Data Release 3.0 (DOI 10.15154/1519007), which includes 3 waves of annual data: baseline (N=11,878), 6 Months (N=11398), Year 1 (N=11,235), 18 Months (N=9911), Year 2 (N=6,571), and 30 Months (N=3601). In terms of race/ethnicity groupings, youth were grouped into non-Hispanic White youth, non-Hispanic Black youth, Black youth, Asian Youth, and Multi-racial race/ethnicity. We used sex at birth, and future work with the ABCD Study will examine gender identity. The demographic distribution of youth in this sample is shown in Table S3.

### Parent Reported Mental Health Symptoms

As described above, parents completed the Child Behavior Checklist at each annual wave. Here we examined raw scores for the following DSM-Oriented symptom subscales: a) total problems; b) internalizing; c) externalizing; d) depression; e) anxiety; f) ADHD; g) oppositional; and h) conduct problems. We used raw scores rather than age and sex-adjusted t-scores to better address relationships to developmental and sex differences.

### Child Reported Mental Health Symptoms

As described above, youth completed the Brief-Problem Monitor (BPM-Y) every six months starting at the first mid-year phone assessment, for a total of 5 waves of assessment (6-Month, Year 1, 18-Month, Year 2, 30-Month). Here we examined raw scores for the four BPM-Y Scales: a) total problems; b) internalizing; c) externalizing; d) attention. We again used raw scores rather than age and sex-adjusted t-scores to better address our questions of interest.

### Neighborhood Poverty

The ABCD Study uses the primary current home address provided by the parent via the Residential History Questionnaire. This address is used to generate the Area Deprivation Index (ADI) for the census tract that contains this address (Singh 2003; Kind and Buckingham 2018). The census data that is used here is from the 2010 Census and some supplementary American Community Survey information. The ADI consists of 17 census variables that use census tracts to reference different aspects of SES. We used the National Percentile metric for the baseline assessment.

### Income-to-needs

Family income at baseline was assessed using the income-to-needs calculated by dividing the reported total household income by the federal poverty line for a given household size (Gonzalez et al. 2020). A higher value indicates higher SES. The gross household income and the number of individuals in the family are reported by the caregiver on the Parent Demographics Survey (Barch et al. 2018).

### Financial Adversity

In addition to Income-to-Needs, we used the Parent-Reported Financial Adversity Questionnaire (PRFQ) (Diemer et al. 2012) assessed at baseline. The PRFQ asked questions designed to determine whether families generally have enough money to pay for basic life expenses like food and healthcare. There are seven questions and each one is scored a 0 or 1. A summary score was created by summing the seven items. PRFQ indexes self-report of finances that may better account for the association of income level to area cost-of-living.

### Caregiver Education

The child’s caregiver reports their education on the Parent Demographics survey. We used the baseline reports and coded 8^th^ grade or lower education as a 1, 9^th^ to 12^th^ without a diploma as a 2, high school, GED or equivalent as a 3, partial college or Associates/vocational degree as a 4, college diploma as a 5, Masters degree as a 6, and a professional or doctorate as a 7.

### Statistical Analysis

We examined trajectories of mental health as a function of age using mixed effects models (MLM’s) in “R” version 4.03 using *lme4* version 1.1.25 for each of the variables that included both random intercept and random slope components (with an unstructured covariance matrix between the two), with nesting in family and site. Time was coded as age at assessments. The models included sex at birth (female, male), race (non-Hispanic White, non-Hispanic Black, Hispanic, Asian, Multi-racial, factor coded with the largest group (White) as the reference group), income-to-needs ratio, financial adversity, caregiver education, and Area Deprivation Index. Maximum likelihood estimation was used. There was missing data for some of the baseline predictors (caregiver education = 23, financial adversity = 22, income-to-needs = 1216, Area Deprivation Index = 697). Thus, we used multiple imputation (N=5) using *mice* version 3.13.0. All continuous predictors were scaled prior to entry into a model using the “scale_datlist” function in miceadds version 3.11-6 to facilitate interpretation of the estimates in terms of effect size. To focus the discussion of effects, and given the large sample size, we employed an ad hoc cutoff of *p* = .01. A full accounting of effects is reported in the text and tables.

## Results

The demographic distribution of the baseline sample is shown in Table S3.

### Brief Problem Monitor-Youth Report

As shown in Table 3 and Figure 1, there were main effects of sex for all measures, as well as interactions of sex with age. To parse the source of these interactions, we created subsets of the imputed data and ran the analyses separately for females and males. As shown in Figure 1, males showed a significant decrease in all symptoms domains with age (all Std. Bs<-0.05, all *ps*<.0086). In contrast, females showed a significant increase in total problems (Std. B=.25, *p*=<.0001), internalizing (Std. B=.17, *p*=<.0001), and externalizing symptoms (Std. B=.05, *p* = .0056) with age, and no significant changes as a function of age in attention problems (Std. B=.05, *p*=.061).

**Table 3:** Results of Analyses of the Brief Problem Monitor Youth (BPM-Y) Report.

|  | **BPM-Y Total Problems** |  |  |  | **BPM-Y Internalizing** |  |  |  |
| --- | --- | --- | --- | --- | --- | --- | --- | --- |
|  | Std. B | 95% CIs | *t* | *p* | Std. B | 95% CI- | *t* | *p* |
| Age (in years) | 0.28 | 0.18 – 0.38 | 5.65 | <.0001 | 0.18 | 0.14 – 0.21 | 8.89 | <.0001 |
| Sex | 0.49 | 0.33 – 0.65 | 5.98 | <.0001 | -0.18 | -0.24 - -0.12 | -5.79 | <.0001 |
| Race/Ethnicity (Factor Coded) |  |  |  |  |  |  |  |  |
| Non-Hispanic Black | 0.07 | -0.22 – 0.37 | 0.50 | 0.6201 | -0.06 | -0.17 – 0.05 | -1.09 | 0.2751 |
| Hispanic | 0.23 | -0.03 – 0.48 | 1.72 | 0.0858 | 0.12 | 0.02 - 0.22 | 2.41 | 0.0159 |
| Asian | -0.37 | -0.95 – 0.22 | -1.23 | 0.2189 | 0.00 | -0.21 – 0.22 | 0.04 | 0.9705 |
| Other | 0.47 | 0.16 – 0.77 | 3.00 | 0.0030 | 0.08 | -0.03 – 0.19 | 1.42 | 0.1554 |
| Caretaker Education | -0.23 | -0.34 - -0.11 | -3.96 | 0.0001 | -0.04 | -0.09 – 0.00 | -2.13 | 0.0340 |
| Income-to-Needs | -0.29 | -0.41 - -0.18 | -4.91 | <.0001 | -0.11 | -0.15 - -0.06 | -4.92 | <.0001 |
| Financial Adversity | 0.48 | 0.39 – 0.57 | 10.22 | <.0001 | 0.14 | 0.11 – 0.18 | 8.34 | <.0001 |
| Area Deprivation Index | 0.23 | 0.10 – 0.36 | 3.46 | 0.0019 | 0.06 | 0.02 – 0.10 | 3.12 | 0.0020 |
| Age X Sex | -0.57 | -0.68 - -0.46 | -10.10 | <.0001 | -0.31 | -0.36 - -0.27 | -13.59 | <.0001 |
| Age X Race/Ethnicity (Factor Coded) |  |  |  |  |  |  |  |  |
| Age X Non-Hispanic Black | -0.52 | -0.72 - -0.32 | -5.14 | <.0001 | -0.24 | -0.32 - -0.16 | -5.88 | <.0001 |
| Age X Hispanic | 0.02 | -0.14 – 0.17 | 0.22 | 0.8260 | -0.04 | -0.11 – 0.02 | -1.41 | 0.1594 |
| Age X Asian | 0.10 | -0.29 – 0.49 | 0.50 | 0.6201 | -0.03 | -0.18 – 0.13 | -0.35 | 0.7290 |
| Age X Other | -0.14 | -0.33 – 0.05 | -1.44 | 0.1507 | -0.04 | -0.11 – 0.04 | -0.91 | 0.3643 |
| Age X Caretaker Education | 0.11 | 0.04 – 0.18 | 3.08 | 0.0021 | 0.07 | 0.04 – 0.10 | 4.91 | <.0001 |
| Age X Income-to-Needs | 0.02 | -0.06 – 0.10 | 0.50 | 0.6208 | 0.01 | -0.03 – 0.04 | 0.38 | 0.7038 |
| Age X Financial Adversity | 0.04 | -0.02 – 0.10 | 1.46 | 0.1452 | 0.02 | 0.00 – 0.05 | 1.62 | 0.1056 |
| Age X Area Deprivation Index | -0.03 | -0.09 – 0.04 | -0.83 | 0.4046 | -0.01 | -0.04 – 0.02 | -0.42 | 0.6774 |
|  | **BPM-Y Externalizing** |  |  |  | **BPM-Y Attention** |  |  |  |
|  | Std. B | 95% CIs | *t* | *p* | Std. B | 95% CI- | *t* | *p* |
| Age (in years) | 0.08 | 0.05 – 0.12 | 4.72 | <.0001 | 0.05 | 0.01 - 0.10 | 2.27 | 0.0233 |
| Sex | 0.24 | 0.19 – 0.30 | 8.17 | <.0001 | 0.44 | 0.36 - 0.52 | 10.83 | <.0001 |
| Race/Ethnicity (Factor Coded) |  |  |  |  |  |  |  |  |
| Non-Hispanic Black | 0.05 | -0.06 – 0.16 | 0.94 | 0.3474 | 0.09 | -0.05 - 0.23 | 1.25 | 0.2124 |
| Hispanic | -0.03 | -0.12 – 0.06 | -0.64 | 0.5240 | 0.17 | 0.05 - 0.29 | 2.73 | 0.0064 |
| Asian | -0.15 | -0.37 – 0.06 | -1.41 | 0.1587 | -0.23 | -0.51 – 0.04 | -1.65 | 0.0990 |
| Other | 0.16 | 0.06 – 0.27 | 3.07 | 0.0022 | 0.29 | 0.16 – 0.43 | 4.22 | <.0001 |
| Caretaker Education | -0.05 | -0.09 - -0.01 | -2.69 | 0.0072 | -0.10 | -0.15 - -0.05 | -3.81 | 0.0002 |
| Income-to-Needs | -0.13 | -0.17 - -0.09 | -6.85 | <.0001 | -0.10 | -0.15 - -0.05 | -3.86 | 0.0001 |
| Financial Adversity | 0.14 | 0.11 – 0.17 | 8.51 | <.0001 | 0.20 | 0.16 – 0.24 | 8.98 | <.0001 |
| Area Deprivation Index | 0.08 | 0.03 – 0.12 | 3.24 | 0.0030 | 0.11 | 0.05 – 0.17 | 3.55 | 0.0011 |
| Age X Sex | -0.16 | -0.20 - -0.12 | -7.66 | <.0001 | -0.14 | -0.20 - -0.09 | -5.45 | <.0001 |

| Age X Race/Ethnicity (Factor Coded) |  |  |  |  |  |  |  |  |
| --- | --- | --- | --- | --- | --- | --- | --- | --- |
| Age X Non-Hispanic Black | -0.07 | -0.14 – 0.00 | -1.96 | 0.0506 | -0.19 | -0.29 - -0.09 | -3.62 | 0.0006 |
| Age X Hispanic | 0.08 | 0.02 – 0.14 | 2.82 | 0.0049 | -0.01 | -0.08 – 0.07 | -0.21 | 0.8322 |
| Age X Asian | 0.04 | -0.10 – 0.17 | 0.52 | 0.6024 | 0.05 | -0.12 – 0.22 | 0.55 | 0.5804 |
| Age X Other | -0.07 | -0.14 - -0.01 | -2.15 | 0.0319 | -0.03 | -0.11 – 0.06 | -0.61 | 0.5390 |
| Age X Caretaker Education | 0.01 | -0.01 – 0.04 | 0.82 | 0.4125 | 0.04 | 0.01 – 0.07 | 2.64 | 0.0082 |
| Age X Income-to-Needs | -0.02 | -0.04 – 0.01 | -1.33 | 0.1846 | 0.02 | -0.01 – 0.05 | 1.19 | 0.2331 |
| Age X Financial Adversity | 0.00 | -0.02 – 0.03 | 0.33 | 0.7441 | 0.03 | 0.00 – 0.06 | 1.94 | 0.0523 |
| Age X Area Deprivation Index | -0.02 | -0.04 – 0.00 | -1.60 | 0.1117 | -0.01 | -0.04 – 0.03 | -0.41 | 0.6822 |

Note: Dark gray indicates effects significant at $p < .01$ , while light gray indicates effects significant at $p < .05$ .

**Figure 1:**
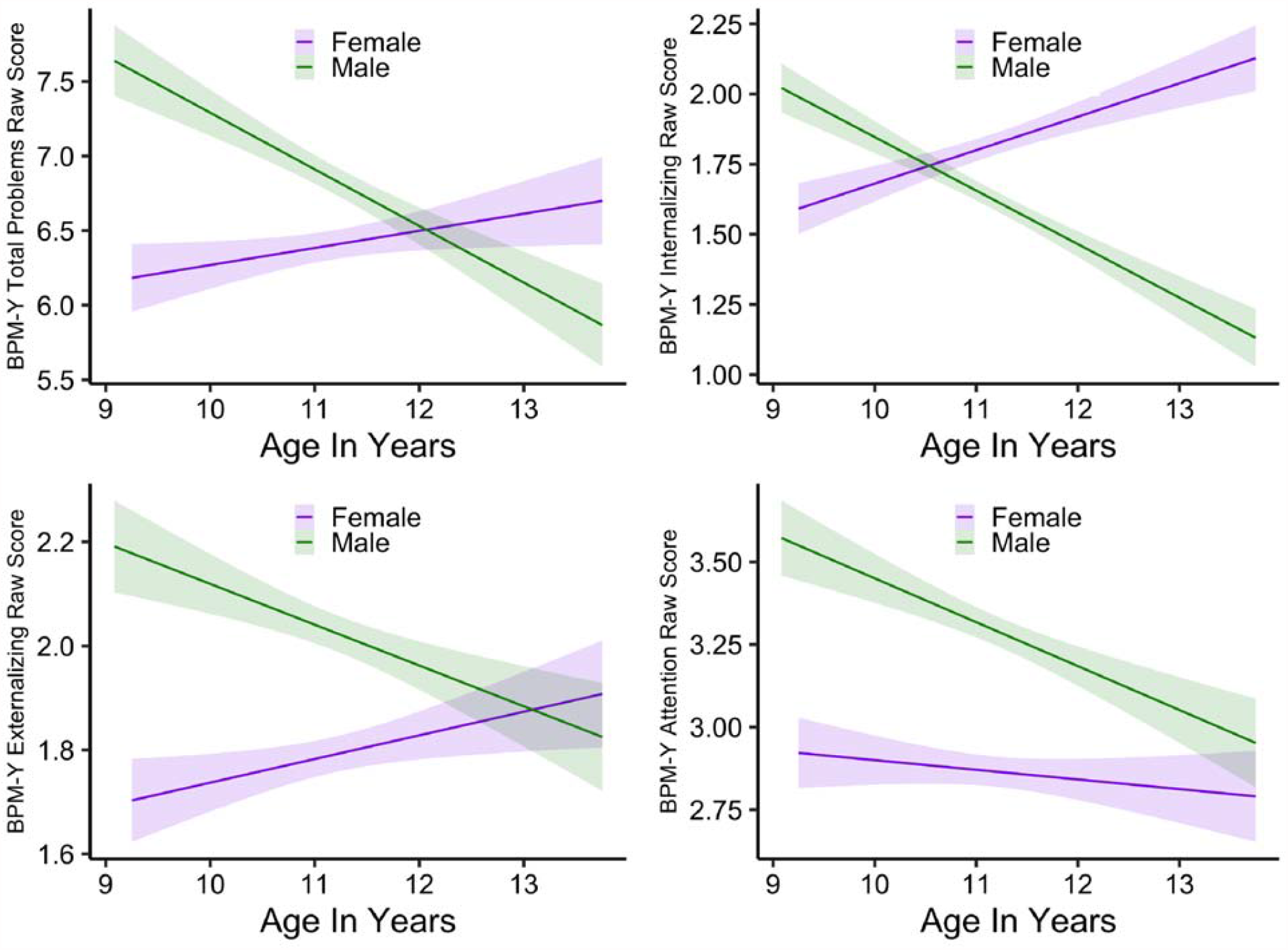
Sex Differences in Trajectories of Youth Reported Mental Health: Graphs illustrating sex differences in youth-reported Total Problem, Internalizing, Externalizing and Attention Problems on the Brief Problem Monitor. The shaded areas indicate the 99^th^ percentile conference intervals around the estimated linear slope. Graphs were created in R using ggplot2 version 3.3.2 using one of the five imputed datasets.

Also as shown in Table 3, there were both main effects of race/ethnicity and interactions with age. As shown in Figure S1, non-Hispanic Black youth did not report overall differences in any problem domain compared to the other race/ethnicity groups, but reported a decrease in total, internalizing, and attention problems with age. The Hispanic youth reported overall higher attention problems compared to non-Hispanic White and Asian youth, and reported a significant increase in externalizing problems with age, both of a small effect size. The other race youth reported overall higher total, externalizing, and attention problems compared to non-Hispanic White and Asian youth. Neither non-Hispanic White or Asian youth reported any mean changes in any problem domain over time (Figure S1).

As shown in Table 3 and Figure S2, there were main effects of caretaker education, income-to-needs, financial adversity, and area deprivation index for every youth reported problem domain (except caretaker education for internalizing problems). These were all simultaneously significant. Higher income-to-needs was associated with lower youth reported problems in all domains, while higher financial adversity and area deprivation were associated with higher youth reported problems in all domains. Higher caretaker education was associated with overall lower youth reported externalizing problems across ages with no significant change as a function of age. Further, there were significant interactions of parent education with age for total, internalizing and attention domains. As shown in Figure S2, while lower caretaker education was associated with greater youth-reported total, internalizing and attentional problems as younger ages, problem reports decreased significantly as youth grew older for those whose caretakers had lower education levels, becoming less elevated compared to the reports of youth with more highly educated caretakers.

### Child Behavior Checklist-Parent Report

There were again main effects of sex and/or interactions of sex with age for the CBCL summary measures (Table 4). As shown in Figure 2, caregivers reported males having overall higher total and externalizing problems. Reports of both total and externalizing problems decreased for both males as females as they grew older, with a steeper decline for males than females. For internalizing problems, males and females were initially rated similarly, but reports increased for females as they moved into puberty, while they decreased for males. There were also main effects of race/ethnicity for all summary measures, with Asian, non-Hispanic Black Youth, and Hispanic youth reported as having lower Total and Externalizing problems compared to Non-Hispanic White youth once SES factors were accounted for (Table 4). There was a similar pattern for internalizing, though it was not significant for Hispanic youth. A significant interaction with age for Non-Hispanic Black youth for internalizing problems also emerged, with a decline in caretaker reported problems with age (Figure S3). Significant interactions of caretaker education with age for both total and externalizing problems were also found. As shown in Figure 3, there were overall lower problem reports for youth among more highly educated caregivers, and less change over time among the youth of caregivers with advanced degrees compared to those with less education. There were main effects of Income-to-Needs for all summary scores, with higher Income-to-Needs associated with lower caregiver reports of problems. There were also main effects and/or interactions with financial adversity for all summary domains. As shown in Figure S4, caretakers with financial adversity reported higher problems for their youth in all domains, with less of a decrease in total problems over time compared to caretakers without financial adversity.

**Table 4:**
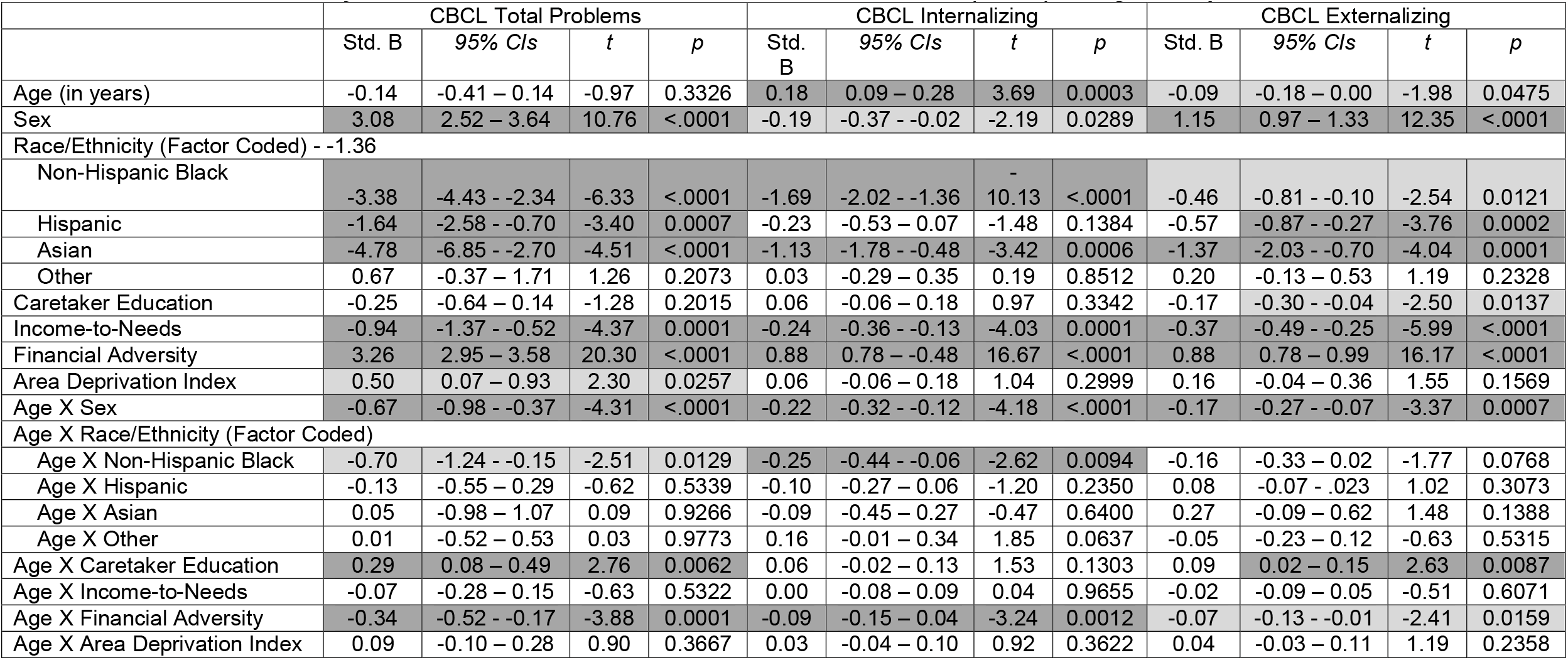
Results of Analyses of the Global Scales for Child Behavior Checklist (CBCL) Caregiver Report.

**Figure 2:**
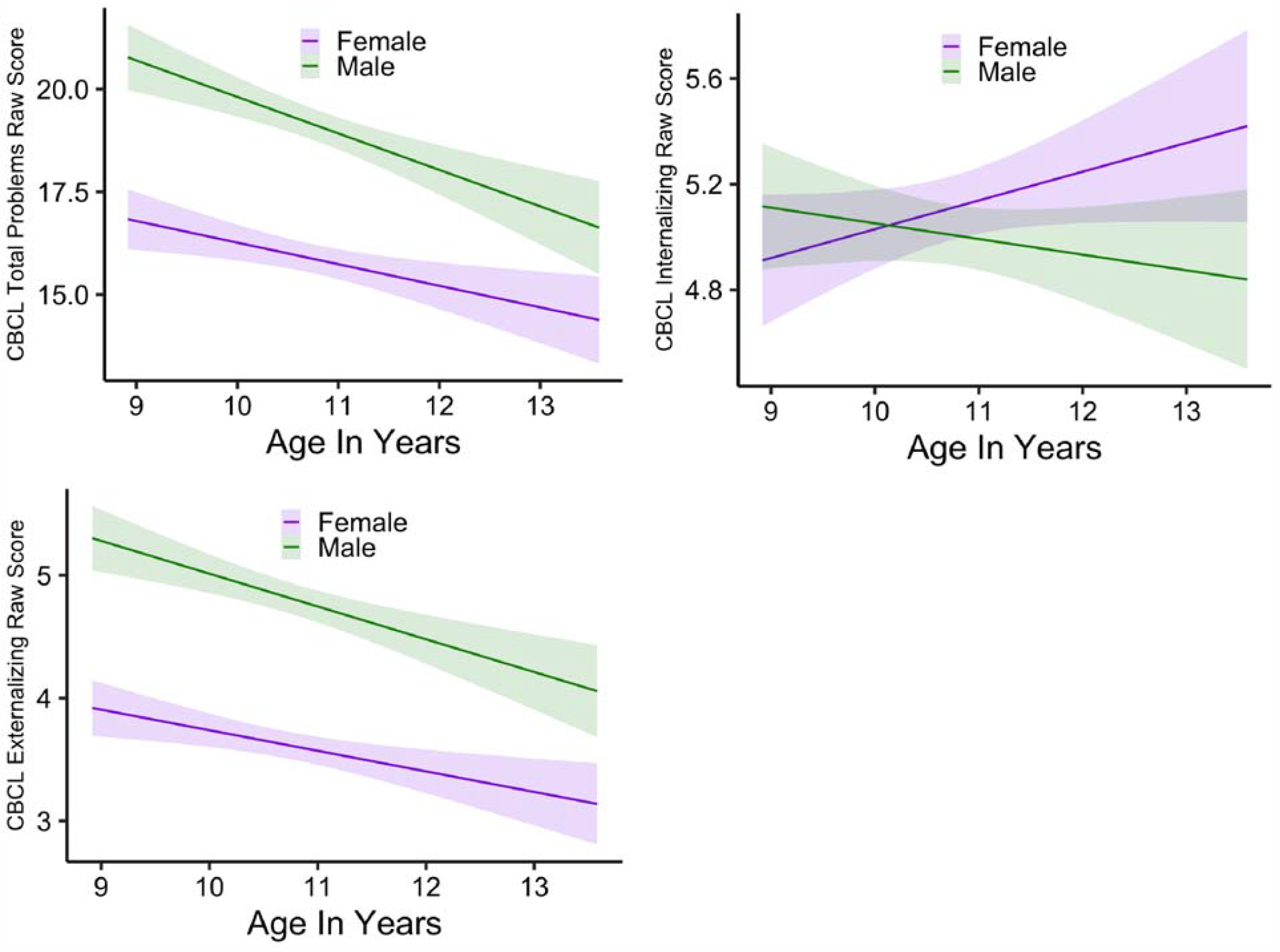
Race/Ethnicity Differences in Trajectories of Caregiver Reported Mental Health Summary Scores: Graphs illustrating race/ethnicity differences in caregiver-reported Total Problem, Internalizing, Externalizing on the Child Behavior Checklist. The shaded areas indicate the 99^th^ percentile conference intervals around the estimated linear slope. Graphs were created in R using ggplot2 version 3.3.2 using one of the five imputed datasets.

**Figure 3:**
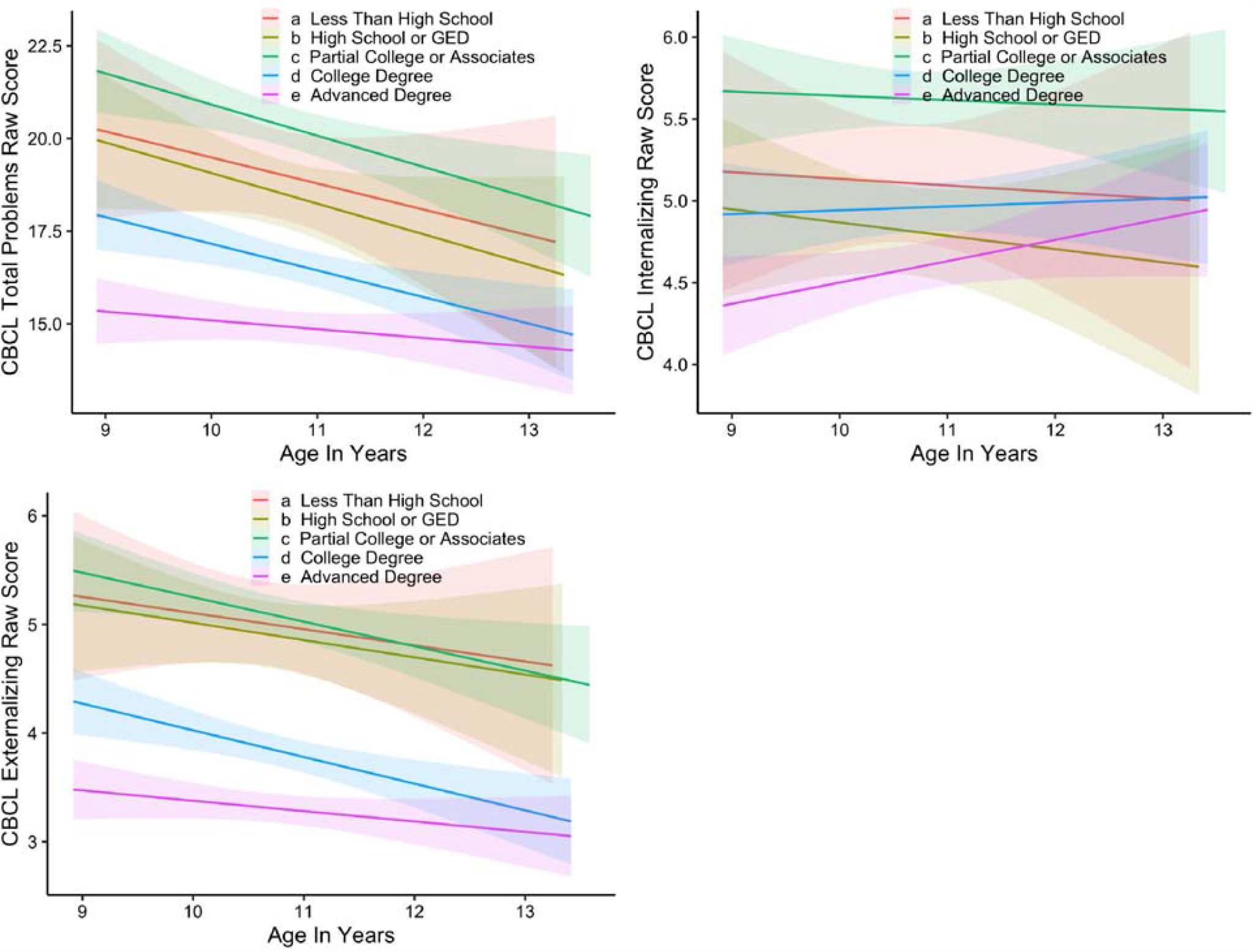
Caretaker Education Differences in Trajectories of Caregiver Reported Mental Health Summary Scores: Graphs illustrating differences in caregiver-reported Total Problem, Internalizing, Externalizing on the Child Behavior Checklist as a function of caretaker education. The shaded areas indicate the 99^th^ percentile conference intervals around the estimated linear slope. Graphs were created in R using ggplot2 version 3.3.2 using one of the five imputed datasets.

Further, caretakers with financial adversity reported an increase in internalizing problems in their youth with age, but not those with no financial adversity. Interestingly, there were no significant unique effects of Area Deprivation Index for caregiver reported mental health, in contrast to youth reported mental health. Results for the DSM Syndrome Scores of depression, anxiety, ADHD, oppositional, and conduct problems are provided in the *Supplemental Materials* (Tables S4 and S5, Figures S5 to S8).

## 13. Discussion of Mental Health Trajectory Results

These analyses of the trajectories of youth and caregiver reported mental health resemble findings from prior work in a number of important ways, but also provide new insights into relationships with socioeconomic disadvantage. In terms of age and sex-related trajectories, like much prior research, we found that both youth and caregiver reports of attentional and externalizing problems were higher for males, but declined with age for both males and females (Bongers et al. 2003; Strohschein 2005; Fanti and Henrich 2010; Robbers et al. 2010) in terms of caregiver report. Intriguingly, in terms of youth report, females actually reported greater externalizing symptoms as they transitioned into puberty, a finding somewhat different than prior work. Also similar to prior research, we found that both youth and caregiver of internalizing symptoms increased with age for females, though not for males (Strohschein 2005; Van Oort et al. 2009; Robbers et al. 2010; Ormel et al. 2012; Ferro et al. 2015; Coley et al. 2019; Antolin-Suarez et al. 2020), with elevated rates for females emerging in adolescence (Bongers et al. 2003; Brown et al. 2007; Robbers et al. 2010; Faravelli et al. 2013). These data add to those in the literature suggesting that the majority of patterns of age and sex-related differences in externalizing and internalizing have maintained over secular changes in society and across a number of different cultures, though with some variation across youth and caregiver report.

Our results in regards to race/ethnicity differences are also intriguing, and suggest relatively few race/ethnicity differences when SES factors are included in models. For youth report, there were only modest race/ethnicity differences, with Multi-racial youth reporting higher total, externalizing and attentional problems, and non-Hispanic Black youth reporting greater decreases in total, internalizing, and attention symptoms than other racial/ethnic groups over time. There were more differences in caregiver-reports, with the caregivers of non-Hispanic White youth reporting higher levels of total, internalizing, and externalizing problems compared to most of the other racial/ethnic groups other than the other-race group when accounting for socio-demographic factors. As noted above, prior findings in regards to race/ethnicity differences in mental health among Black, Asian and Hispanic youth have been somewhat mixed, but our findings are consistent with a number of studies showing lower rates of internalizing symptoms among non-Hispanic Black youth (Coley et al. 2019) and findings that attention symptoms were higher among White youth (Collins and Cleary 2016). However, we would still caution any strong interpretations of these data, as we were not able to address a range of other potentially contextualizing factors, such as cultural differences in reporting of mental health related symptoms, or experiences of discrimination or other forms of systemic racism that may influence mental health.

Consistent with the robust previous literature on SES and mental health, we found that all indicators of lower SES were related to greater total, internalizing, and externalizing problems in youth reports. Critically, other than a few exceptions, all SES indicators accounted for simultaneous unique variance in youth mental-health reports, including income-to-needs, caretaker education, financial adversity, and area deprivation index. For caregiver reports, all SES indicators other than Area Deprivation Index also accounted for unique variance in reports of youth mental health, though caregiver education effects were less consistent than income-to-needs and financial adversity. It is intriguing that neighborhood poverty has stronger effects on youth reported mental health than caregiver reported mental health. It is unclear why this was so, but it is possible that youth’s experiences with their peers, in their schools, and in their neighborhoods are likely to be reflected in their own introspective reports than in their caregiver’s. Overall, these data once again indicate the huge importance of SES in understanding youth mental health, and indicate the need to understand the many different facets of SES given that they each account for independent variance in the trajectory of youth mental health.

Of note, all of these data were collected prior to the onset of the COVID-19 pandemic, and thus it will be important in future analyses to examine discontinuities that may occur following the onset of the pandemic, and how individual differences in pre-pandemic factors predict response to the pandemic restrictions or to COVID-19 infection itself. In particular, it will be important to examine how various facets of SES related to the impact of COVID-19, and whether youth who were showing particular trajectories of change in mental health (e.g., greater than average increase in internalizing, etc.) are more likely to have been negatively impacted by facets of the pandemic. Sadly, COVID-19 may also create an experiment of nature by which we are able to examine the relationships of changes in SES to changes in youth mental health, as the SES of many ABCD families, like so many families across the world, was negatively impacted by COVID-19.

## 14. Summary and Conclusions

This manuscript outlined the logic and known issues with the measures of mental health that have been included in the ABCD Study since baseline and which have been added over time. Further, we have tried to outline known issues to alert the field to some of the challenges with certain measures, as well as the promise afforded by this rich, multi-informant, longitudinal database. We believe that the analyses of the longitudinal trajectories of both youth and caregiver reported mental health illustrate the power of this data set in identifying the ways in which mental health evolves in children as a function of a variety of factors, including age, sex, and SES. As additional waves of ABCD data are released, we will be able to examine additional factors that may impact mental health among youth in the United States, and began to examine leading and lagging relationships of risk factors for increasing mental health challenges, resilience factors that may protect some youth, and the consequences of changes in mental health over time.

## Data Availability

All of the data are publically available through the National Data Archive

https://nda.nih.gov/abcd/query/abcd-curated-annual-release-3.0.html

## Acknowledgements

We wish to thank all of the wonderful families who participate in the ABCD Study, and all of the outstanding staff who make this work possible. We also wish to thank Drs. Ken Kobak and Dr. Joan Kaufman for their support of the KSADs assessments and Dr. Thomas Achenbach for his support for the CBCL, BPM, ASR, ABCL, and YSR assessments. We also wish to thank Dr. Marybel Robledo Gonzalez for sharing her computation of income-to-needs ratios for use in our analyses.

Data used in the preparation of this article were obtained from the Adolescent Brain Cognitive Development (ABCD) Study (https://abcdstudy.org), held in the NIMH Data Archive (NDA). This is a multisite, longitudinal study designed to recruit more than 10,000 children age 9-10 and follow them over 10 years into early adulthood. The ABCD Study is supported by the National Institutes of Health and additional federal partners under award numbers U01DA041022, U01DA041028, U01DA041048,U01DA041089, U01DA041106, U01DA041117, U01DA041120, U01DA041134, U01DA041148,U01DA041156, U01DA041174, U24DA041123, and U24DA041147. A full list of supporters is available at https://abcdstudy.org/nih-collaborators. A listing of participating sites and a complete listing of the study investigators can be found at https://abcdstudy.org/principal-investigators.html.ABCD consortium investigators designed and implemented the study and/or provided data but did not necessarily participate in analysis or writing of this report. This manuscript reflects the views of the authors and may not reflect the opinions or views of the NIH or ABCD consortium investigators. The ABCD data repository grows and changes over time. The ABCD data used in this report came from DOI: 10.15154/1519007. DOIs can be found at https://nda.nih.gov/abcd/query/abcd-curated-annual-release-3.0.html.

