## Supplemental Material for "Special Issue: Demographic and Mental Health Assessments in the Adolescent Brain and Cognitive Development Study: Updates and Longitudinal Trajectories"

**Table S1: Members of the ABCD Mental Health Assessment Workgroup as of April, 2021**

| **Name** | **Role** | **Institution** |
| --- | --- | --- |
| Deanna M. Barch | Co-Chair | Washington University in St. Louis |
| Alexandra Potter | Co-Chair | University of Vermont |
| Mathew Albaugh | Member | University of Vermont |
| Shelli Avenevoli | Member | National Institute of Mental Health |
| Arielle Basking-Sommers | Member | Yale University |
| Dara Blachman-Demner | Member | Office of Behavioral and Social Sciences Research |
| Brittany Bryant | Member | Medical University of South Carolina |
| Duncan Clark | Member | University of Pittsburgh Medical School |
| John Foxe | Member | University of Rochester |
| Dylan Gee | Member | Yale University |
| Jay Giedd | Member | University of California at San Diego |
| Meyer Glantz | Member | National Institute on Drug Abuse |
| James J. Hudziak | Member | University of Vermont |
| Michelle Johns | Member | Center for Disease Control and Prevention |
| Nicole Karcher | Member | Washington University in St. Louis |
| Barbara Kelley | Member | National Institute of Justice |
| Christine Larson | Member | University of Wisconsin-Milwaukee |
| Kimberly LeBlanc | Member | National Institute of Drug Abuse |
| Melanie Maddox | Trainee Member | University of Utah |
| Erin McGlade | Member | University of Utah |
| Carrie Mulford | Member | National Institute of Drug Abuse |
| Bonnie Nagel | Member | Oregon Health & Science University |
| Gretchen Neigh | Member | Virginia Commonwealth University |
| Clare Palmer | Post-Doctoral Member | University of California at San Diego |
| Kenneth Sher | Member | University of Missouri at Columbia |
| Susan Tapert | Member | University of California at San Diego |
| Laili Xie | Trainee Member | University of California at San Diego |

**Table S2: KSADS-COMP Modules Administered at Each Wave**

|  |  | **Baseline** | | **Year 1** | | **Year 2** | | **Year 3** | | **Year 4** | | **Year 5** | | **Year 6#** | |
| --- | --- | --- | --- | --- | --- | --- | --- | --- | --- | --- | --- | --- | --- | --- | --- |
|  |  | **Age 9-10** | | **Age 10-11** | | **Age 11-12** | | **Age 12-13** | | **Age 13-14** | | **Age 14-15** | | **Age 15-16** | |
| Module # | Module Name | Y | P | Y | P | Y | P | Y | P | Y | P | Y | P | Y | P |
| 1-3 | Mood Disorders | 1 | 1 | 0 | 0 | 1 | 1 | 0 | 0 | 1 | 1 | 0 | 0 | 1 | ? |
| 4 | Psychosis | 0 | 1 | 0 | 1 | 0 | 1 | 0 | 1 | 0 | 1 | 0 | 1 | ? | 1 |
| 5 | Panic Disorder | 0 | 1 | 0 | 0 | 0 | 1 | 0 | 0 | 0 | 1 | 0 | 0 | ? | 1 |
| 6 | Agoraphobia | 0 | 1 | 0 | 0 | 0 | 1 | 0 | 0 | 0 | 1 | 0 | 0 | ? | 1 |
| 7 | Separation Anxiety | 0 | 1 | 0 | 0 | 0 | 1 | 0 | 0 | 0 | 0 | 0 | 0 | 0 | 0 |
| 8 | Social Anxiety Disorder | 1 | 1 | 0 | 0 | 1 | 1 | 0 | 0 | 1 | 1 | 0 | 0 | 1 | 1 |
| 9 | Specific Phobia | 0 | 1 | 0 | 0 | 0 | 1 | 0 | 0 | 0 | 1 | 0 | 0 | ? | 1 |
| 10 | Generalized Anxiety Disorder | 1 | 1 | 0 | 0 | 1 | 1 | 0 | 0 | 1 | 1 | 0 | 0 | 1 | 1 |
| 11 | Obsessive Compulsive Disorder | 0 | 1 | 0 | 0 | 0 | 1 | 0 | 0 | 0 | 1 | 0 | 0 | 1 | 1 |
| 12 | Enuresis and Encopresis | 0 | 0 | 0 | 0 | 0 | 0 | 0 | 0 | 0 | 0 | 0 | 0 | 0 | 0 |
| 13 | Eating Disorders | 0 | 1 | 0 | 1 | 1 | 1 | 0 | 1 | 1 | 1 | 0 | 1 | 1 | 1 |
| 14 | Attention Deficit Hyperactivity Disorder | 0 | 1 | 0 | 1 | 0 | 1 | 0 | 1 | 0 | 1 | 0 | 1 | ? | 1 |
| 15 | Oppositional Defiant Disorder | 0 | 1 | 0 | 1 | 0 | 1 | 0 | 0 | 0 | 1 | 0 | 0 | 0 | 0 |
| 16 | Conduct Disorder | 0 | 1 | 0 | 1 | 1 | 1 | 0 | 1 | 1 | 1 | 0 | 1 | 1 | 1 |
| 17 | Tic Disorders | 0 | 0 | 0 | 0 | 0 | 0 | 0 | 1 | 0 | 1 | 0 | 1 | 0 | ? |
| 18 | Autism Spectrum Disorders | 0 | 1 | 0 | 0 | 0 | 1 | 0 | 0 | 0 | 1 | 0 | 0 | 0 | 1 |
| 19 | Alcohol Use Disorder | 0 | 1 | 1* | 0 | 1* | 1 | 1* | 0 | 1* | 1 | 1* | 0 | 1* | 1 |
| 20 | Drug Use Disorders | 0 | 1 | 1* | 0 | 1* | 1 | 1* | 0 | 1* | 1 | 1* | 0 | 1* | 1 |
| 21 | Post-Traumatic Stress Disorder | 0 | 1 | 0 | 0 | 0 | 1 | 0 | 0 | 0 | 1 | 0 | 0 | ? | 1 |
| 22 | Sleep Problems | 1 | 1 | 0 | 0 | 1 | 1 | 0 | 0 | 1 | 1 | 0 | 0 | 1 | 1 |
| 23 | Suicidality | 1 | 1 | 1 | 0 | 1 | 1 | 1 | 0 | 1 | 1 | 1 | 0 | 1 | 1 |
| 24 | Homicidality | 0 | 1 | 0 | 0 | 0 | 1 | 0 | 0 | 0 | 1 | 0 | 0 | ? | 1 |
| 25 | Selective Mutism | 0 | 0 | 0 | 0 | 0 | 0 | 0 | 0 | 0 | 0 | 0 | 0 | 0 | 0 |

Note: Y = Youth; P = Parent; *Only administered if youth reported substance use. #Year six is not yet confirmed or piloted and is subject to change.

**Table S3: Participant Characteristics**

|  | **Total**  **N** | **N** | **Percent** |  |  |  |  |
| --- | --- | --- | --- | --- | --- | --- | --- |
| Female sex at birth | 11878 | 5682 | 47.8% |  |  |  |  |
| Race/Ethnicity | 11878 |  |  |  |  |  |  |
| Non-Hispanic White |  | 6181 | 52.0% |  |  |  |  |
| Non-Hispanic Black |  | 1784 | 15.0% |  |  |  |  |
| Hispanic |  | 2412 | 20.3% |  |  |  |  |
| Asian |  | 252 | 2.1% |  |  |  |  |
| Multiracial |  | 1248 | 10.5% |  |  |  |  |
| Caretaker Eduation | 11855 |  |  |  |  |  |  |
| Less Than High School |  | 785 | 6.6% |  |  |  |  |
| High School or GED |  | 1258 | 10.6% |  |  |  |  |
| Partial College or Associates Degree |  | 3486 | 29.4% |  |  |  |  |
| College Degree |  | 3331 | 28.1% |  |  |  |  |
| Advanced Degree |  | 2995 | 25.3% |  |  |  |  |
|  | **N** | **Mean** | **SD** | **Min** | **Max** | **Skew** | **Kurtosis** |
| Baseline Income-to-Needs | 10662 | 39.44 | 28.61 | 0.33 | 153.9 | 0.734 | -0.09 |
| Baseline Financial Aversity | 11856 | 0.47 | 1.10 | 0 | 7 | 2.81 | 8.32 |
| Baseline Area Deprivation Index | 11181 | 39.24 | 27.3 | 0 | 100 | 0.64 | -0.54 |

**Table S4: Results of Analyses of the Depression and Anxiety Scales for Child Behavior Checklist Caregiver Report**

|  | CBCL Depression | | | CBCL Anxiety | | |
| --- | --- | --- | --- | --- | --- | --- |
|  | Std. B | *t* | *p* | Std. B | *t* | *p* |
| Age (in years) | 0.18 | 9.50 | <.0001 | -0.01 | -0.57 | 0.5704 |
| Sex | 0.16 | 4.69 | <.0001 | -0.08 | -2.13 | 0.0329 |
| Race/Ethnicity (Factor Coded) | | | | | | |
| Non-Hispanic Black | -0.54 | -8.66 | <.0001 | -0.67 | -8.98 | <.0001 |
| Hispanic | -0.12 | -2.19 | 0.0289 | -0.07 | -1.00 | 0.3170 |
| Asian | -0.39 | -3.16 | 0.0016 | -0.51 | -3.52 | 0.0004 |
| Other | 0.01 | 0.11 | 0.9154 | 0.00 | -0.01 | 0.9922 |
| Caretaker Education | 0.01 | 0.23 | 0.8203 | 0.06 | 2.14 | 0.0330 |
| Income-to-Needs | -0.09 | -3.75 | 0.0002 | -0.11 | -4.14 | <.0001 |
| Financial Adversity | 0.35 | 18.40 | <.0001 | 0.29 | 12.50 | <.0001 |
| Area Deprivation Index | 0.02 | 0.96 | 0.3381 | 0.00 | 0.02 | 0.9837 |
| Age X Sex | -0.54 | -8.66 | <.0001 | -0.08 | -3.55 | 0.0004 |
| Age X Race/Ethnicity (Factor Coded) | | | | | | |
| Age X Non-Hispanic Black | -0.07 | -3.29 | 0.0011 | -0.08 | -1.87 | 0.0647 |
| Age X Hispanic | -0.13 | -3.41 | 0.0008 | -0.09 | -2.57 | 0.0105 |
| Age X Asian | -0.04 | -1.41 | 0.1591 | -0.05 | -0.62 | 0.5385 |
| Age X Other | -0.11 | -1.39 | 0.1659 | 0.03 | 0.61 | 0.5411 |
| Age X Caretaker Education | 0.05 | 1.34 | 0.1839 | 0.02 | 1.64 | 0.1012 |
| Age X Income-to-Needs | 0.02 | 1.11 | 0.2686 | 0.00 | -0.10 | 0.9213 |
| Age X Financial Adversity | -0.01 | -0.52 | 0.6052 | -0.04 | -3.14 | 0.0020 |
| Age X Area Deprivation Index | -0.01 | -0.66 | 0.5088 | -0.01 | -0.63 | 0.5337 |

**Table S5: Results of Analyses of the ADHD, Oppositional, and Conduct Sub-Scales for Child Behavior Checklist Caregiver Report**

|  | CBCL ADHD | | | CBCL Oppositional | | | CBCL Conduct | | |
| --- | --- | --- | --- | --- | --- | --- | --- | --- | --- |
|  | Std. B | *t* | *p* | Std. B | *t* | *p* | Std. B | *t* | *p* |
| Age (in years) | -0.07 | -3.16 | 0.0016 | -0.05 | -2.79 | 0.0053 | -0.01 | -0.36 | 0.7209 |
| Sex | 0.91 | 18.84 | <.0001 | 0.39 | 12.03 | <.0001 | 0.53 | 14.29 | <.0001 |
| Race/Ethnicity (Factor Coded) | | | | | | | | | |
| Non-Hispanic Black | -0.15 | -1.75 | 0.0796 | -0.34 | -5.67 | <.0001 | 0.08 | 1.27 | 0.2046 |
| Hispanic | -0.19 | -2.35 | 0.0191 | -0.18 | -3.43 | 0.0006 | -0.26 | -4.41 | <.0001 |
| Asian | -0.78 | -4.50 | <.0001 | -0.64 | -5.42 | <.0001 | -0.28 | -2.10 | 0.0358 |
| Other | 0.23 | 2.65 | 0.0081 | -0.02 | -0.27 | 0.7852 | 0.19 | 2.87 | 0.0041 |
| Caretaker Education | -0.02 | -0.62 | 0.5357 | -0.01 | -0.22 | 0.8222 | -0.11 | -4.29 | <.0001 |
| Income-to-Needs | -0.08 | -2.36 | 0.0212 | -0.12 | -5.08 | <.0001 | -0.13 | -4.99 | <.0001 |
| Financial Adversity | 0.40 | 13.99 | <.0001 | 0.25 | 13.89 | <.0001 | 0.32 | 15.55 | <.0001 |
| Area Deprivation Index | 0.08 | 2.26 | 0.0262 | 0.03 | 1.10 | 0.2730 | 0.09 | 3.10 | 0.0039 |
| Age X Sex | -0.06 | -2.26 | 0.0255 | -0.05 | -2.68 | 0.0073 | -0.06 | -2.60 | 0.0095 |
| Age X Race/Ethnicity (Factor Coded) | | | | | | | | | |
| Age X Non-Hispanic Black | -0.12 | -2.62 | 0.0090 | 0.01 | 0.41 | 0.6852 | -0.09 | -2.34 | 0.0194 |
| Age X Hispanic | -0.07 | -1.78 | 0.0782 | 0.01 | 0.42 | 0.6740 | 0.04 | 1.38 | 0.1703 |
| Age X Asian | 0.01 | 0.13 | 0.8968 | 0.08 | 1.33 | 0.1853 | 0.09 | 1.23 | 0.2190 |
| Age X Other | -0.06 | -1.38 | 0.1688 | 0.00 | -0.02 | 0.9835 | -0.01 | -0.27 | 0.7893 |
| Age X Caretaker Education | 0.03 | 1.83 | 0.0674 | 0.02 | 1.27 | 0.2071 | 0.03 | 2.00 | 0.0509 |
| Age X Income-to-Needs | -0.01 | -0.72 | 0.4702 | 0.00 | -0.38 | 0.7077 | -0.02 | -1.13 | 0.2609 |
| Age X Financial Adversity | -0.03 | -1.89 | 0.0599 | -0.01 | -1.30 | 0.1955 | -0.02 | -1.42 | 0.1600 |
| Age X Area Deprivation Index | 0.01 | 0.45 | 0.6584 | 0.02 | 1.21 | 0.2319 | 0.00 | 0.28 | 0.7789 |

**Figure Captions**

**Figure S1: Race/Ethnicity Differences in Trajectories of Youth Reported Mental Health**: Graphs illustrating race/ethnicity differences in youth-reported Total Problem, Internalizing, Externalizing and Attention Problems on the Brief Problem Monitor. The shaded areas indicate the 99^th^ percentile conference intervals around the estimated linear slope. Graphs were created in R using ggplot2 version 3.3.2 using one of the five imputed datasets.

**Figure S1: Caretaker Education Differences in Trajectories of Youth Reported Mental Health**: Graphs illustrating differences in youth-reported Total Problem, Internalizing, Externalizing and Attention Problems on the Brief Problem Monitor as a function of caretaker education level. The shaded areas indicate the 99^th^ percentile conference intervals around the estimated linear slope. Graphs were created in R using ggplot2 version 3.3.2 using one of the five imputed datasets.

**Figure S3: Race/Ethnicity Differences in Trajectories of Caregiver Reported Mental Health Summary Scores**: Graphs illustrating race/ethnicity differences in caregiver-reported Total Problem, Internalizing, Externalizing on the Child Behavior Check List . The shaded areas indicate the 99^th^ percentile conference intervals around the estimated linear slope. Graphs were created in R using ggplot2 version 3.3.2 using one of the five imputed datasets.

**Figure S4: Financial Adversity Differences in Trajectories of Caregiver Reported Mental Health Summary Scores**: Graphs illustrating differences in caregiver-reported Total Problem, Internalizing, Externalizing on the Child Behavior Check List as a function of caretaker reported financial adversity. Scores of 0 on this measure were coded as no adversity, and scores of 1+ were coded as adversity for the purposes of illustration. The shaded areas indicate the 99^th^ percentile conference intervals around the estimated linear slope. Graphs were created in R using ggplot2 version 3.3.2 using one of the five imputed datasets.

**Figure S5: Sex Differences in Trajectories of Caregiver Reported Mental Health DSM Syndrome Scores**: Graphs illustrating sex differences in caregiver-reported Depression, Anxiety, ADHD, Oppositional, and Conduct scores on the Child Behavior Check List . The shaded areas indicate the 99^th^ percentile conference intervals around the estimated linear slope. Graphs were created in R using ggplot2 version 3.3.2 using one of the five imputed datasets.

**Figure S6: Race/Ethnicity Differences in Trajectories of Caregiver Reported Mental Health DSM Syndrome Scores**: Graphs illustrating race/ethnicity differences in caregiver-reported Depression, Anxiety, ADHD, Oppositional, and Conduct scores on the Child Behavior Check List . The shaded areas indicate the 99^th^ percentile conference intervals around the estimated linear slope. Graphs were created in R using ggplot2 version 3.3.2 using one of the five imputed datasets.

**Figure S7: Caretaker Education Differences in Trajectories of Caregiver Reported Mental Health DSM Syndrome Scores**: Graphs illustrating differences in caregiver-reported Depression, Anxiety, ADHD, Oppositional, and Conduct scores on the Child Behavior Check List as a function of caretaker education level. Scores of 0 on this measure were coded as no adversity, and scores of 1+ were coded as adversity for the purposes of illustration. The shaded areas indicate the 99^th^ percentile conference intervals around the estimated linear slope. Graphs were created in R using ggplot2 version 3.3.2 using one of the five imputed datasets.

**Figure S8: Financial Adversity Differences in Trajectories of Caregiver Reported Mental Health DSM Syndrome Scores**:: Graphs illustrating differences in caregiver-reported Depression, Anxiety, ADHD, Oppositional, and Conduct scores on the Child Behavior Check List as a function of caretaker reported financial adversity. Scores of 0 on this measure were coded as no adversity, and scores of 1+ were coded as adversity for the purposes of illustration. The shaded areas indicate the 99^th^ percentile conference intervals around the estimated linear slope. Graphs were created in R using ggplot2 version 3.3.2 using one of the five imputed datasets.
