## Supplementary figures and images for "Special Issue: Demographic and Mental Health Assessments in the Adolescent Brain and Cognitive Development Study: Updates and Longitudinal Trajectories"

### Supplemental Figures

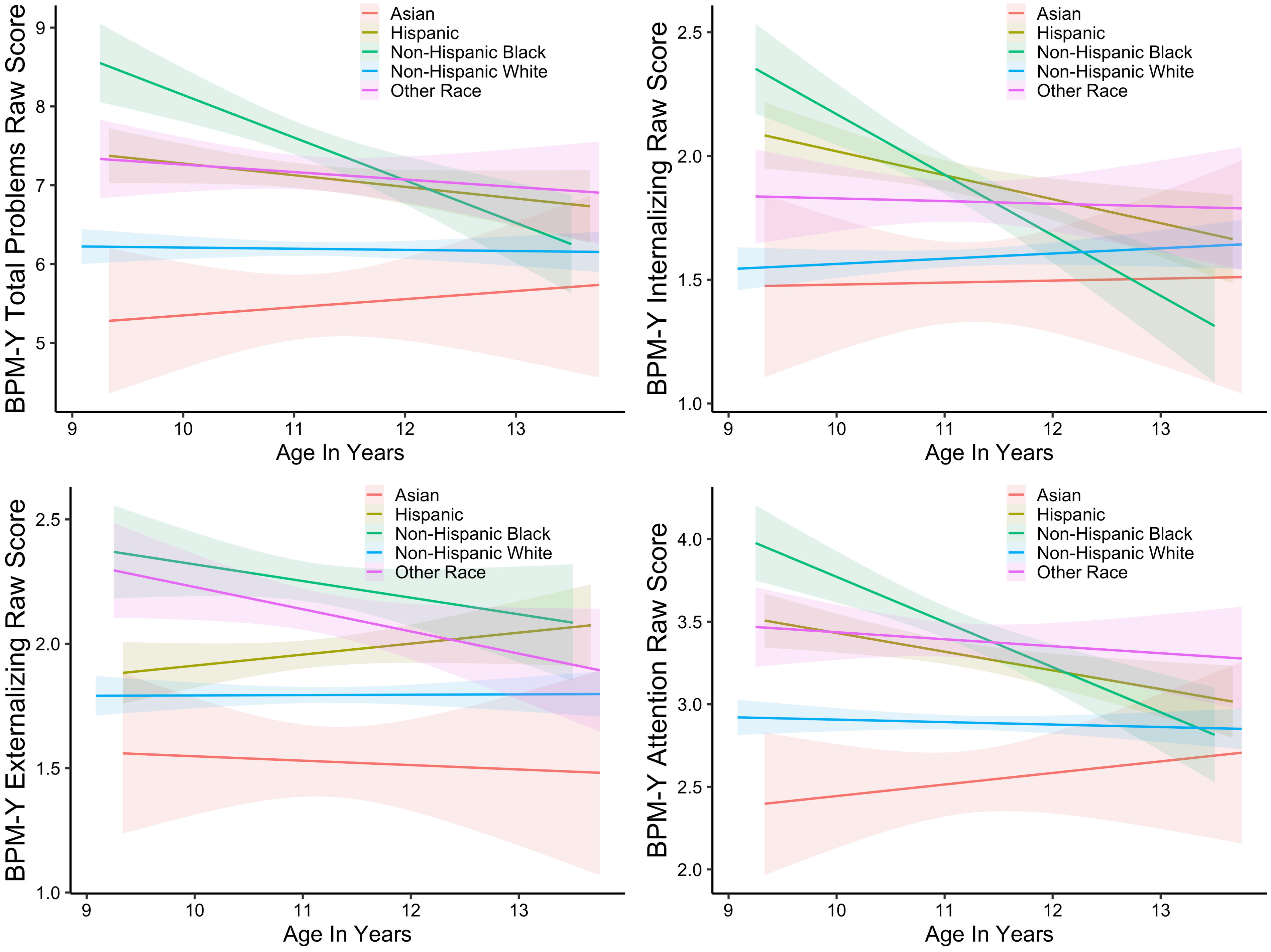

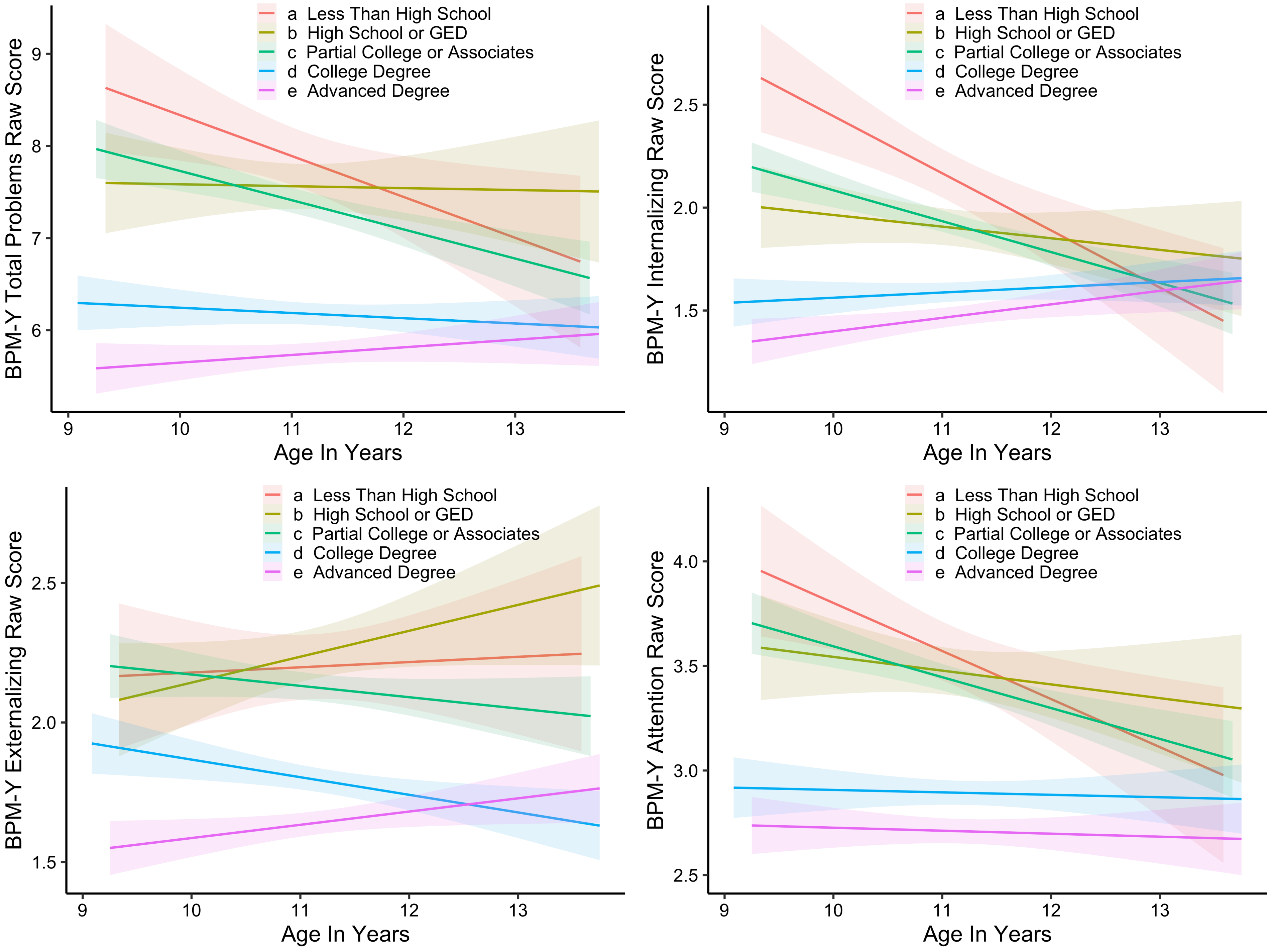

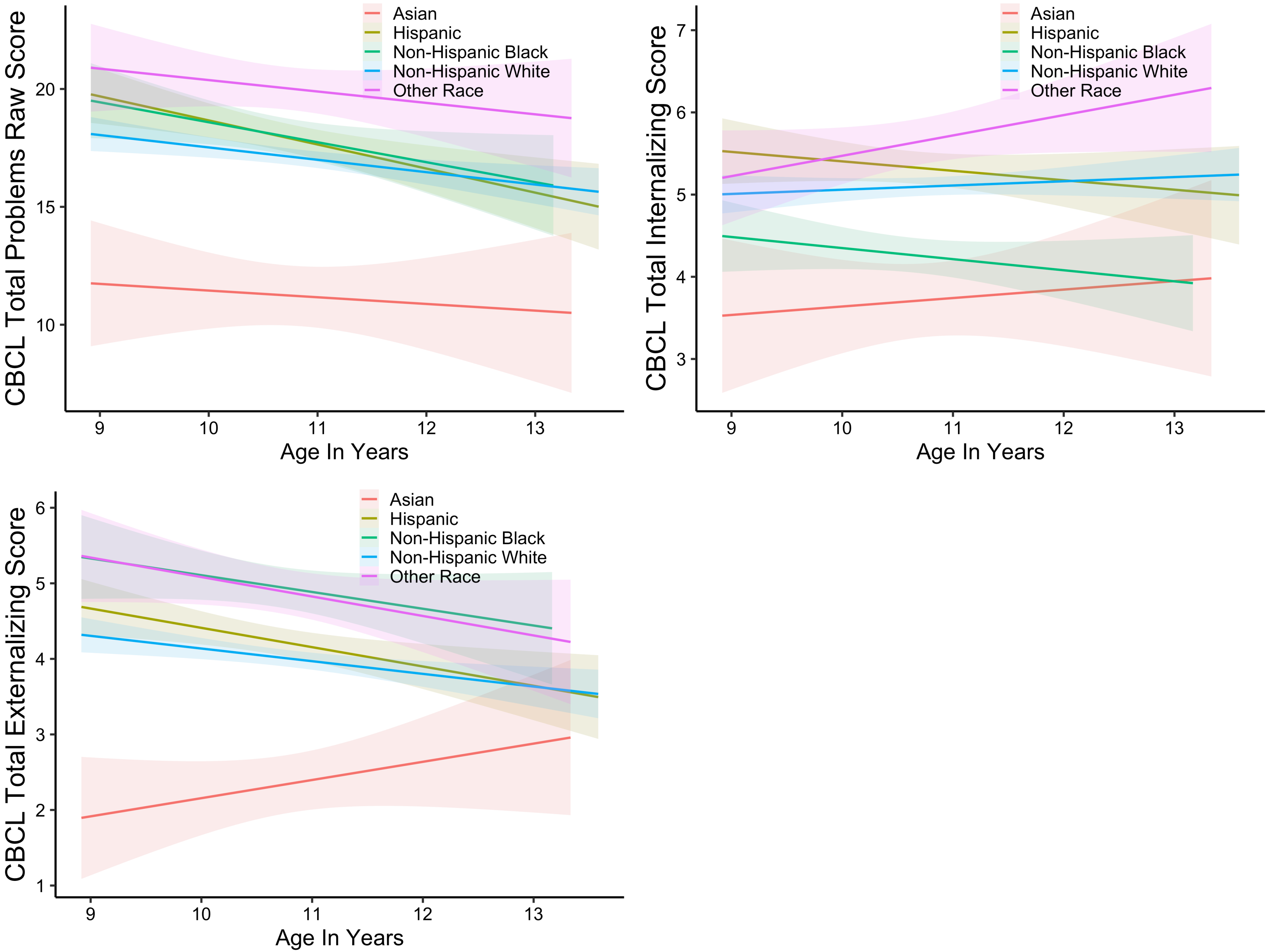

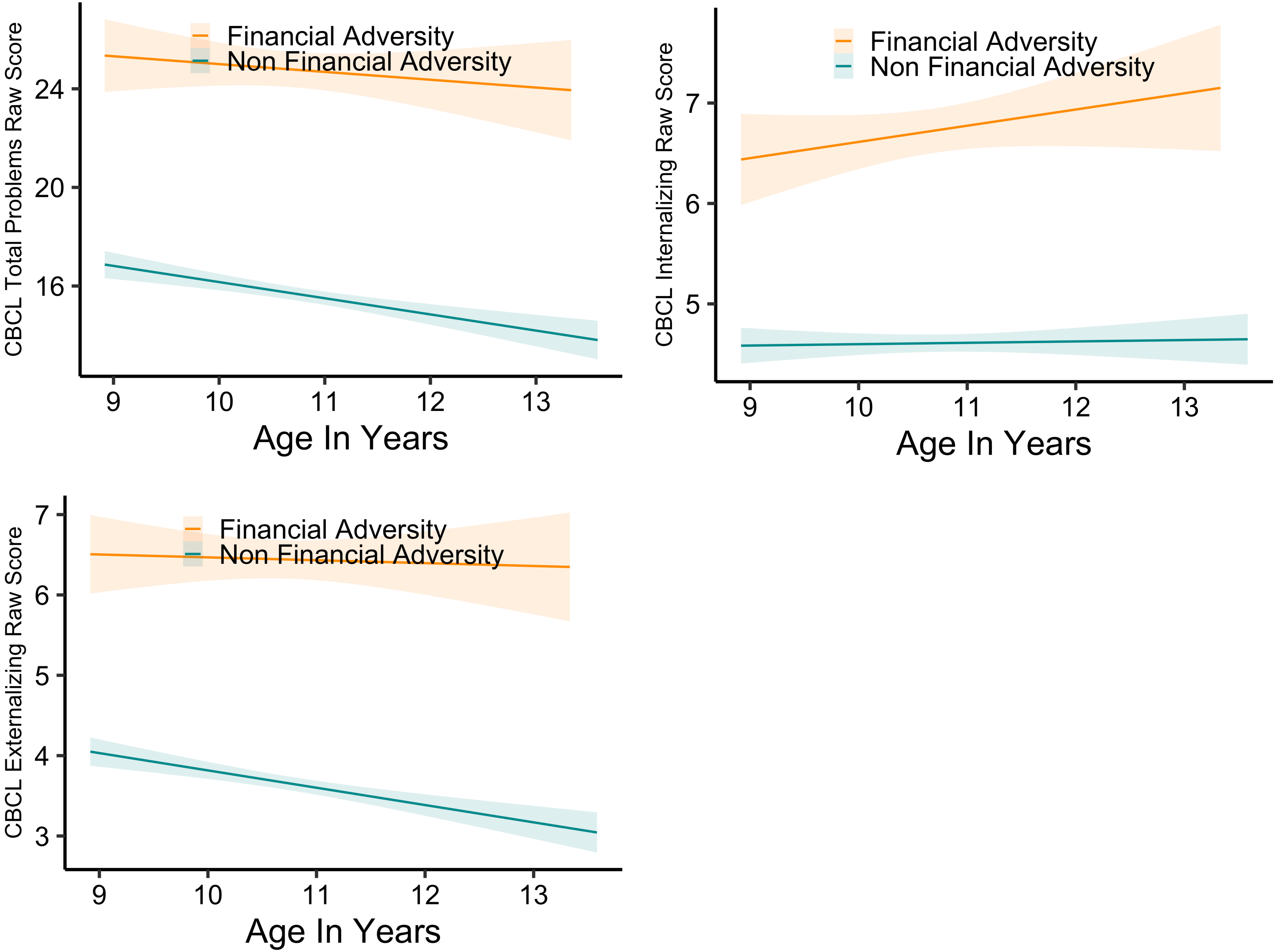

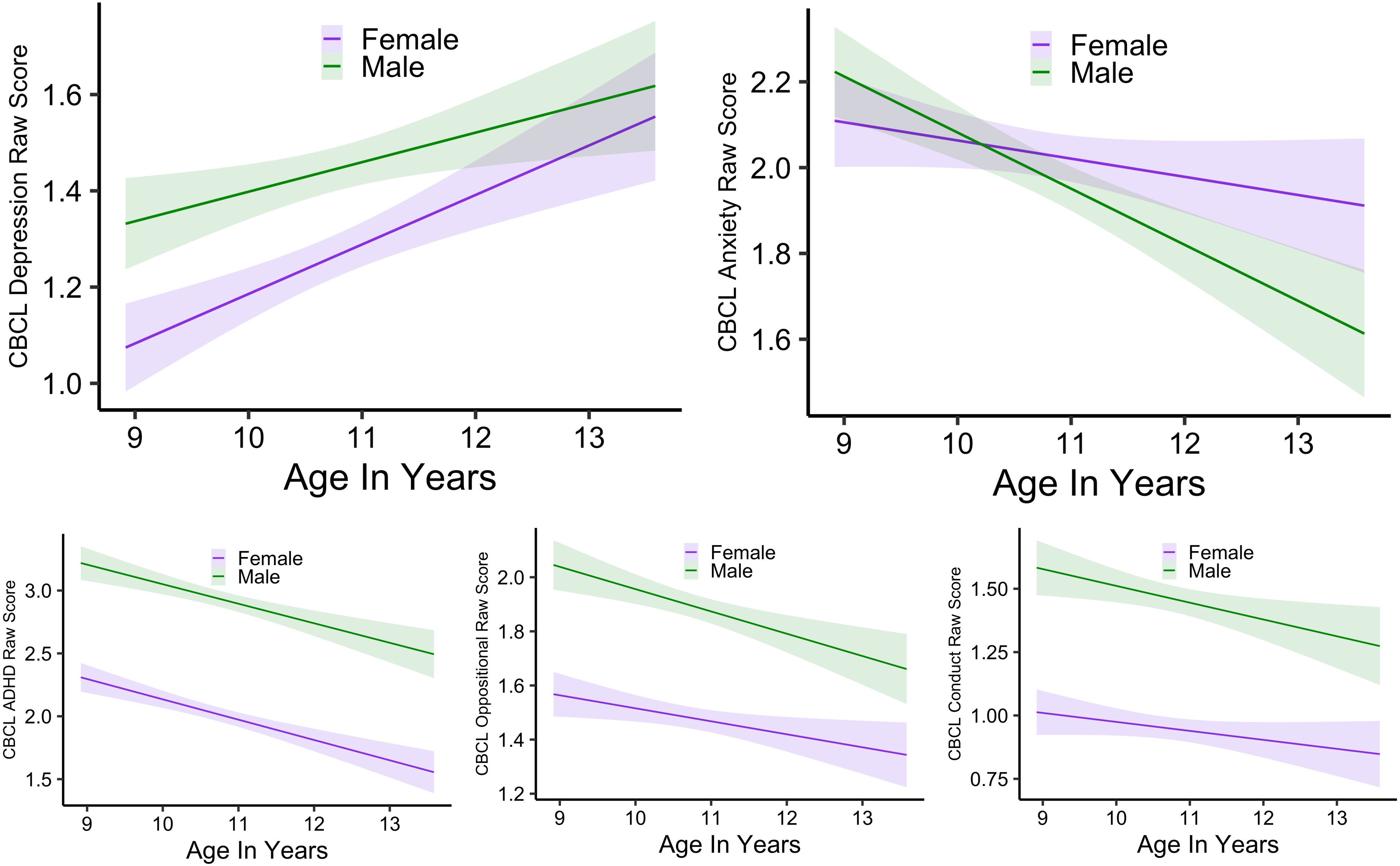

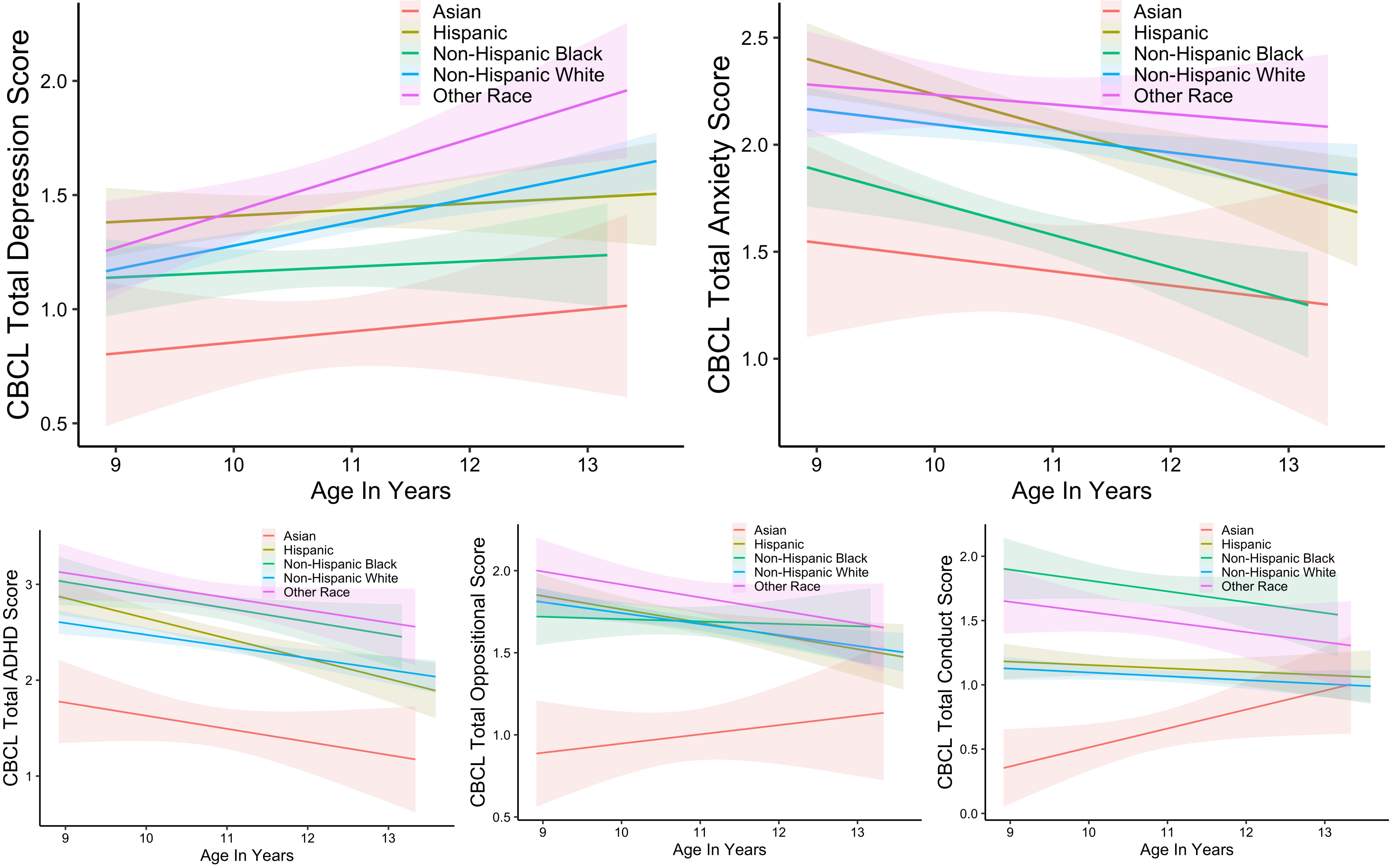

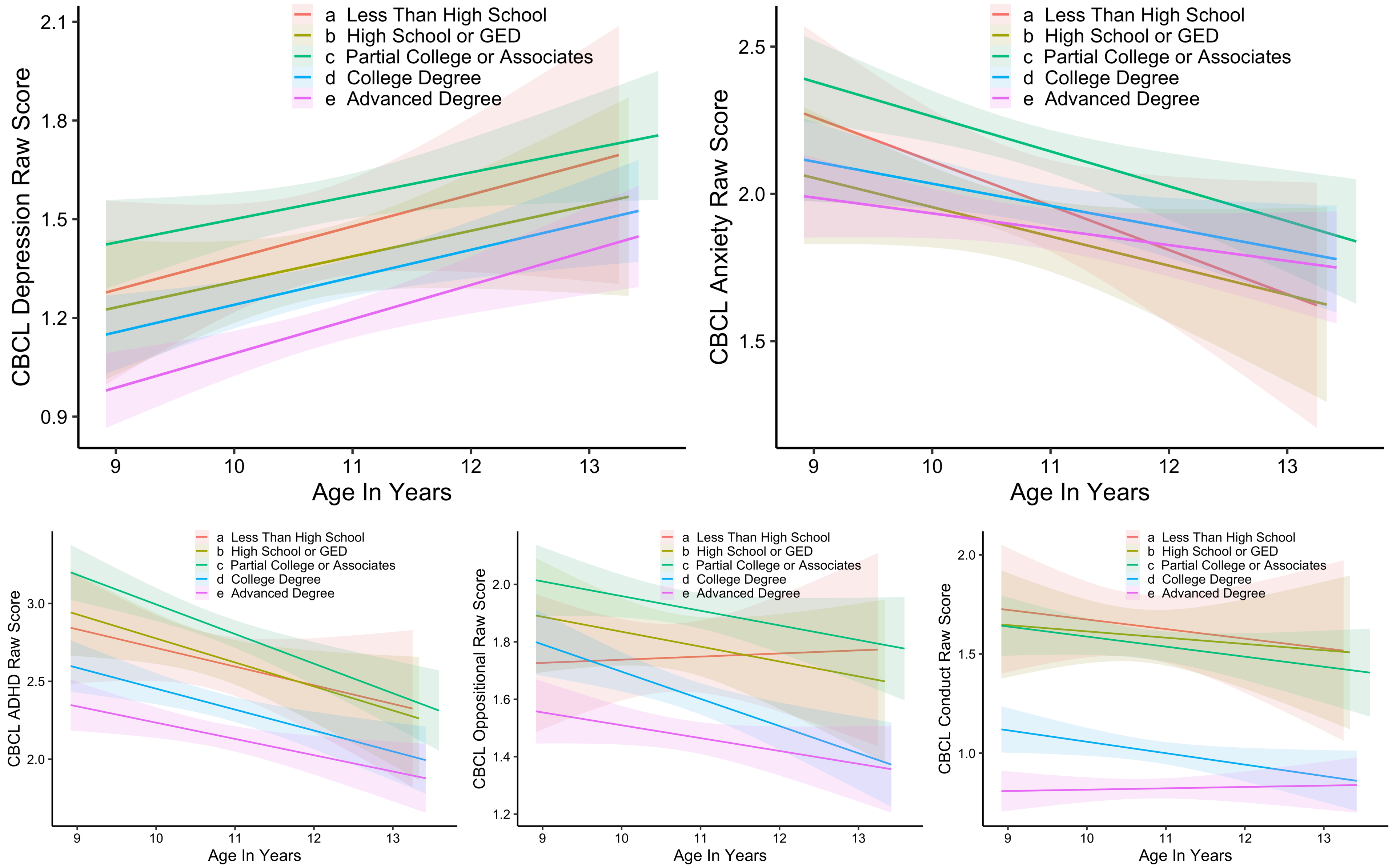

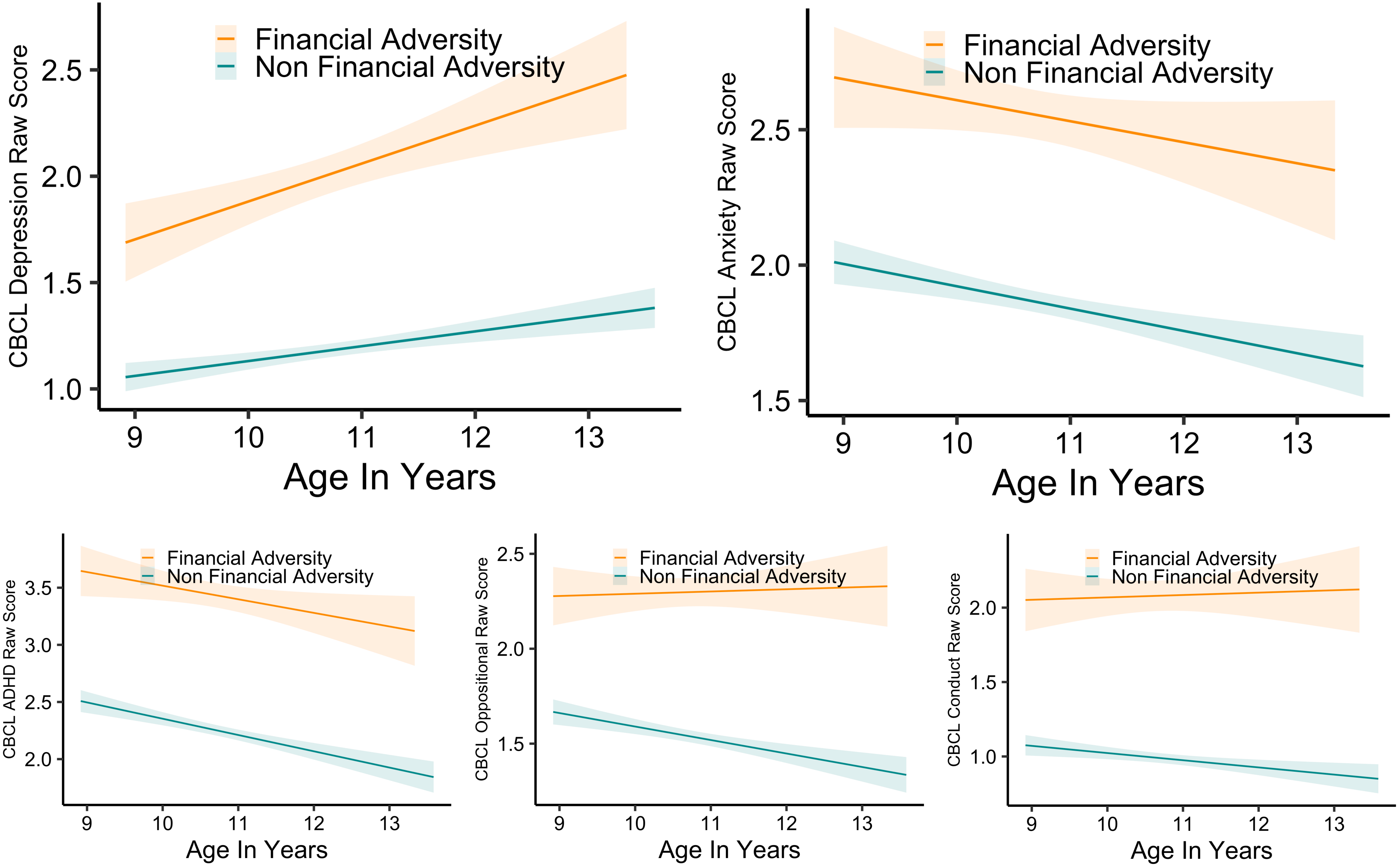
